# Genotypic and phenotypic analysis of 173 patients with extremely rare pathogenic mutations who applied for experimental antisense oligonucleotide treatment

**DOI:** 10.1101/2024.08.05.24310862

**Authors:** Stanley T. Crooke, Tracy Cole, Jeffrey B. Carroll, Joseph G. Gleeson, Laurence Mignon, Julie Douville, Wendy Chung, Jennifer Bain, Elizabeth Berry-Kravis, Nelson Leung, Andy Watt, Berit Powers, Cedrik Tekendo-Ngongang, Stanislav Kmoch, Petr Vyletal, Sarah Glass

## Abstract

Recent advances in “omics” technologies allow for the identification of an increasing number of individuals with diseases caused by nano-rare mutations. These difficult-to-diagnose individuals are uniquely disadvantaged and pose significant challenges to healthcare systems and society. Despite having diseases caused by actionable single gene mutations, in many cases, there is no commercial path for treatments for such small patient populations. We have defined nano-rare mutations as, mutations with a known worldwide prevalence <30. Since antisense oligonucleotide (ASO) technology has proven to be suited to address the needs of a portion of these patients, the n-Lorem Foundation is establishing an industrialized approach that couples detailed genotypic and phenotypic data to the immediate potential for ASO therapy. In this manuscript we have leveraged our experience in assessing the causality of nano-rare genetic variants and associated proximal molecular pathological events to attempt a correlation between detailed genetic data with patient specific phenotypic observations in 173 nano-rare individuals from diverse age groups evaluated for experimental ASO therapy. We found that the time required to achieve a molecular diagnosis varies from 1 month to 36 years, with the mean and median times from symptom onset to diagnosis estimated to be 4.32 years and 2 years, respectively. Amongst submitted cases there is a significant bias toward neurological diseases, with diverse genes and functional families involved and a marked preponderance of mutations in ion channel genes. The variability in phenotypic expression associated with nano-rare variants in genes such as *GNAO1, H3F3A, GBE1, UBTF,* or *PACS1* clearly supports previous observations that phenotypes associated with same variants in the same gene can vary. We also observe that different, but functionally equivalent variants can result in both similar (e.g., *TARDBP*) and different phenotypes (e.g., *GNAO1*). Despite the relatively small size of the patient population investigated, this first compilation of its kind allows a variety of insights into the genotype and phenotype relationships in nano-rare conditions. Moreover, we show that our unique patient population presents a remarkable opportunity to apply “modern omics” approaches to begin to understand the various homeostatic, compensatory, and secondary effects of these genetic variants on the networks that result in expression of their unique phenotypes.

To provide a more detailed description of the processes involved to provide a personalized antisense medicine, we have included nonclinical and clinical data on three exemplary patients who display disease in three different organs, the CNS, the eye and the kidney and are treated with ASOs of different designs. In contrast to traditional drug development, each patient presents unique genomic, ASO design, clinical treatment and management and evaluation challenges.

## INTRODUCTION

Advances in genetic sequencing, transcriptomics, single cell RNA sequencing and a variety of other “omics” and the publication of multiple genome-wide association studies (GWAS) databases have transformed biomedical research. Moreover, these databases continue to evolve with the addition of new information[1–3]. Basic insights include the estimate that the average human genome has tens of million single nucleotide polymorphisms (SNPs) distributed non-randomly across and within various genes. There are also multiple types of mutations in addition to SNPs, and a fraction of these mutations are associated with pathology. The proliferation of multiple genomic databases and the continued expansion of genomic information provide excellent tools for cross-species comparisons, phenotype-genome and disease-genome relationships and sub- population comparisons[4]. Despite this broad progress, there are few, if any, databases that can link genomic and other “omic” characteristics to the phenotypes of specific patients[5–7]. Without such linkages, it is difficult to correlate individual patient genomics and phenotypes to large population studies. Nor is it possible to couple genomic and phenotypic characteristics with traditional prospective epidemiological studies[8]. Further, only by coupling genomic and phenotypic data, is it possible to consider personalized treatments for individual patients. Clearly, multiple publications have compared various genotypes with phenotypes in specific diseases, such as Prader-Willi syndrome ^9^ and intellectual disability ^10^and these provide important insights. However, an analysis unbiased by disease that considers any phenotype associated with a specific genotype adds a mutation focused assessment that may be valuable. Our mutation-directed research effort is creating such a database for extremely rare mutations that is orthogonal to traditional disease-centered approaches.

Patients with diseases caused by extremely rare mutations pose particularly difficult challenges to today’s healthcare systems [9]. Nano-rare patients are defined as individuals with genetic diseases caused by mutations with a known worldwide prevalence of <30 [9, 10]. Though the precise prevalence of nano-rare mutations is unknown and will remain unknown until population-wide genomic sequencing of newborns is implemented, based on estimates derived from the Undiagnosed Disease Network (UDN), these patients may number in the millions [11]. According to the UDN, it is estimated that the average time from symptom onset to diagnosis is 8 years for patients with difficult to diagnose diseases and it is estimated that the vast majority of nano-rare patients are never diagnosed. However, no systematic study of nano-rare patients that correlates detailed genotypic characteristics with phenotypic characteristics has been reported [12, 13]. As nano-rare patients represent the rarest of mutations, a database such as ours may provide important insights that enable a better understanding of this very minimally studied group of patients.

Our focus is to provide treatments to patients with pathogenic nano-rare mutations, which are, by definition, the rarest of genetic diseases, because patients with these mutations are uniquely disadvantaged. Our focus also is advantageous as an adduct to other databases that focus on genes or diseases because it is orthogonal to other approaches. Many, if not most, nano-rare patients have syndromes that have not previously been identified and, based on our initial experience, some patients are truly unique, i.e., they have mutations that are novel to them. Further, nano-rare patients who have a previously identified syndrome typically harbor an extremely rare mutation that may result in different signs and symptoms from the more common causal mutation. These patients are isolated and often unable to identify an expert who can make a diagnosis or provide even a rudimentary prognosis. In the search for a diagnosis, nano-rare patients are typically mis- referred, mis-diagnosed and mis-treated while the disease progresses. Moreover, at least today, there is no commercial path for treatments for patient populations this small. However, nano-rare patients have one distinct advantage: they have diseases caused by identifiable and understandable mutations in single genes (monogenic).

n-Lorem is a non-profit foundation initiated in January 2020 with the mission of taking advantage of antisense technology to discover, develop, manufacture and provide experimental antisense oligonucleotide (ASO) treatments to appropriate nano-rare patients for free for life[12]. Importantly, we have established a clear path from case submission to n-Lorem through treatment of accepted patients (Supplemental Fig. 1). Our goal is to industrialize the processes that result in the exposure of nano-rare patients to experimental ASOs. This means that each step that leads to the selection of an ASO for each patient must be of the highest quality possible and the clinical exposure of each patient must be managed professionally by individuals experienced in conducting clinical trials. It is also vital that those responsible for managing patients exposed to personalized ASOs be aware of clinical results reported for other ASOs of the same chemical class, especially with a focus on any novel safety observations.

To learn maximally from each patient and our aggregate experience, we have adapted the traditional crossover design employed in single patient studies^11,12^. Both because our patients are severely ill and ASOs have long durations of action, we work with the clinical investigator to define primary, secondary and exploratory treatment goals and clinical measures that we will use to assess the performance of each ASO in each patient at the initiation of the ASO discovery program. We then collect data focused on these treatment goals during the 12 months or so prior to treatment and compare those analyses on treatment to pre-treatment. This approach, which is described in more detail in the supplementary methods for exemplary patients, has provided solid information with which to judge ASO performance. Importantly, this process has proven to be readily scalable and applicable to all the organs and syndromes that are represented in the first 173 patients.

Antisense technology is ideally suited to address the needs of a meaningful fraction of nano-rare patients. Given that ASOs are genetic medicines, once the causative mutation is identified, its molecular pathologic effects characterized, and the patient’s phenotype defined, ASO discovery efforts for that patient can begin. As most nano-rare patients experience a long odyssey to diagnosis during which their disease progresses, the efficiency and rapidity of ASO discovery are critical. The technology is versatile because multiple post-RNA binding mechanisms have been identified and validated [14–19]. Arguably, the greatest advantage of ASOs is that, within a chemical class, all of these medicines are similar as they differ only in sequence. Thus, optimized ASOs share pharmacokinetic (PK) and pharmacodynamic (PD) properties such as distribution to and within organs and cells, distribution half-life, elimination half-life, potency, and types of adverse events. The aforementioned properties are the subject of scores of primary manuscripts and reviews, including databases that integrate safety observations from all controlled clinical trials for the broadly used chemical classes of ASOs [20–23].

Three major chemical classes of ASOs have been thoroughly studied in humans. Phosphorothioate (PS) 2′methoxyethyl (MOE) ASOs have been evaluated in controlled clinical studies in many thousands of patients. Similarly, PS 2′-MOE N-Acetylgalactosamine (GalNAc) ASOs that enhance productive delivery to hepatocytes have been studied in tens of thousands of patients. Finally, PS 2′ constrained ethyl (cEt) ASOs and their GalNAc counterparts have been evaluated in hundreds of patients. Other chemical classes that have been evaluated in clinical trials include PS locked nucleic acids (LNA) ^26^ and morpholinos (PMO) ^27^Though PS LNA ASOs are quite potent *in vitro* in pre-clinical and clinical trials they have proven to have significant toxicities that are difficult to manage. Consequently, no PS LNA ASOs are approved for commercial use. PMO ASOs are neutral and therefore distribute to peripheral tissues poorly and require very high systemic doses. Nevertheless, they have demonstrated benefit in a single disease, Duchenne muscular dystrophy ^27^. PS, MOE, cEt and LNA ASOs can be designed to take advantage of a wide variety of post-binding mechanisms ^28^. The most frequently used mechanism is designed to cause degradation of target RNAs by RNase H1. Those ASOs are typically referred to as “gapmers” in which the 5′ and 3′ wings incorporate 2′ modifications (e.g. 2′MOE, cEt or LNA) and the center 8 -10 nucleotides are PS deoxynucleotides. For PS ASOs designed to block or enhance translation or alter splicing, the PS ASOs are typically fully 2′ modified, i.e. all 2′ positions contain modifications such as 2′MOE. Being fully modified, when the PS ASOs bind to target RNAs, they do not serve as substrates for RNase H1^16,17^.

Cytotoxicity, innate immune activation, complement activation and overall protein binding are influenced by sequence. Fortunately, the mechanism for cytotoxicity is now well understood ^29^ and, therefore, avoidable. Similarly, the effect of PS ASOs on complement activation is known to be due to binding to factor H, a complement inhibitor ^30^ and is avoidable. However, the structure activity relationships (SAR) of innate immune activation via TLR9 are complex reflecting a lack of full understanding of the full molecular mechanisms ^31-33^. Consequently, to avoid innate immune activation and other potential target effects, AI-informed ASO screening is necessary. We have a proprietary algorithm that is the product of experience with tens of millions of ASOs studied in a wide variety of systems, including data from hundreds of thousands of humans and it evolves steadily as we study new ASOs, generate hypotheses to explain the screening data and incorporate new factors as needed. As the quality of any algorithm is defined by the extent of the database contributing to the algorithm, this experience is valuable. However, the algorithm simply prevents certain ASO sequences from being synthesized. If no algorithm is available, if the steps, number of ASOs at each step and the pass/fail criteria shown in our manuscript are followed, safe and effective ASOs can be identified.

The PS content is the primary determinant of distribution of PS ASOs and the 2′modifications enhance stability and duration of effect. Given that within each chemical class, ASOs differ only by sequence, they share similar PK and PD properties making it possible to pre-define the appropriate route, approximate therapeutic dose, dose frequency and potential adverse events^11,12,14^. Sequence-specific cytotoxicity can be observed with some PS ASO sequences and the mechanism is well understood ^29^. To avoid potentially cytotoxic sequences, we evaluate 400- 500 PS ASO sequences at high does in patient derived cells and standard laboratory cell lines. Some PS ASO sequences can also induce innate immune activation and to avoid such ASOs all are screened in the BJAB cell assay system that has been shown to identify innate immune activating PS ASOs ^34^. Potent and well-tolerated PS ASOs are then subjected to 8-week tolerability screens in rodents and the best performing PS ASO is evaluated in a 3-month good laboratory practice (GLP) toxicology study. The specific numbers of ASOs and the pass/fail criteria for each step are shown in Table 1. At each step in the process, only PS ASOs that meet the pass/fail criteria for that step proceed to the next step in the evaluation. However, even with the rapidity and efficiency of ASO technology, it would not be possible to initiate a personalized ASO discovery program and hope to initiate dosing of a patient within 15-18 months were it not for the special guidance of well understood chemical classes, issued by the FDA^12,35,36^ (Supplemental Fig. 2).

**Table 1:**
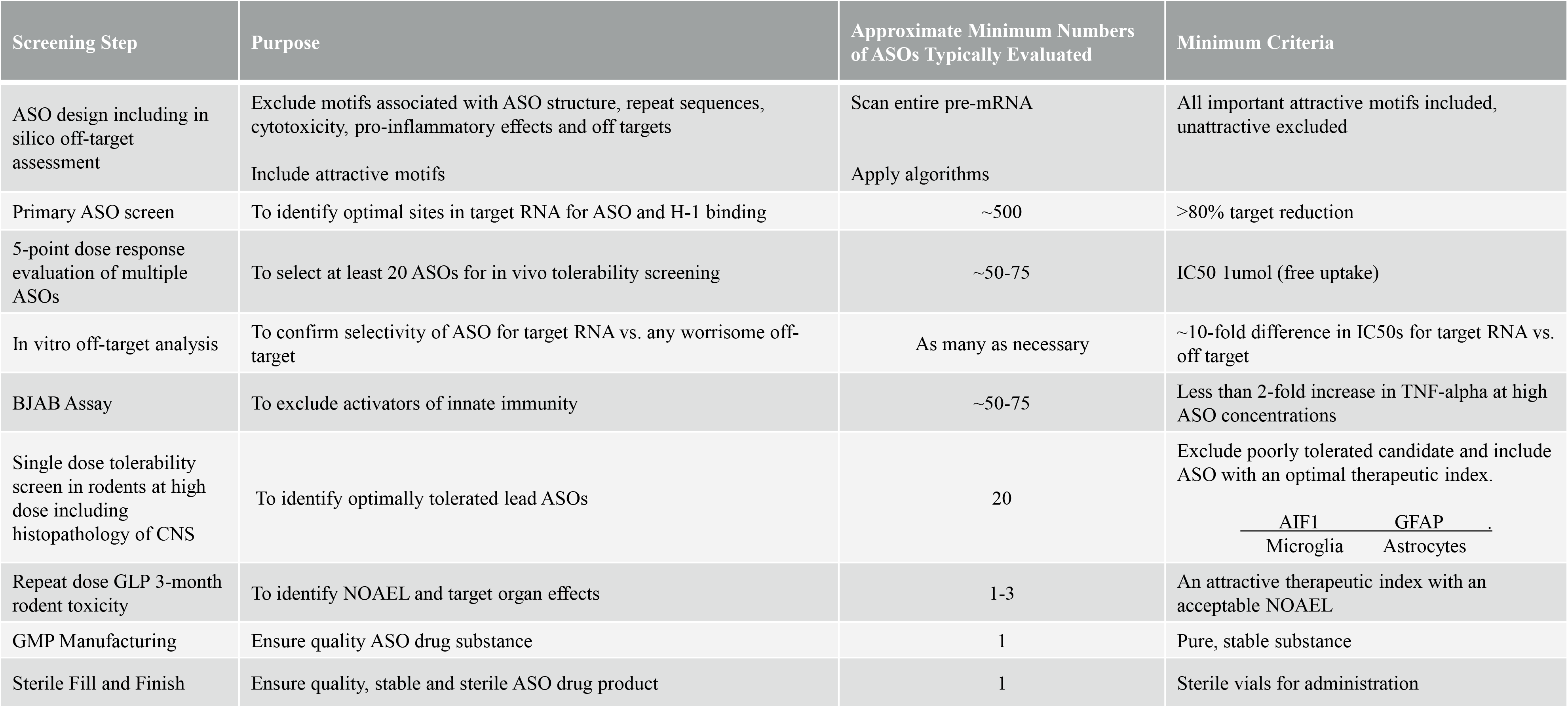
Current n-Lorem Process To Discover And Develop Optimal RNase H1 ASOs for CNS Diseases.

After processing 173 individual patient applications, we performed a data cut-off and analyzed this body of information. The purposes of this manuscript are to determine which patients are amenable to ASO treatment, establish ASO discovery and development processes, and define methods used to evaluate the clinical effects of the ASOs on individual patients. Importantly, the characteristics of this, the largest cohort of patients nano-rare mutations reported to date and the results of ASO treatment of some of these patients will be reported. To provide added granularity, we summarize the nonclinical and clinical data from three exemplary patients with nano-rare mutations that manifest in different organs.

## METHODS

### FDA nonclinical guidance

The FDA’s nonclinical guidance for personalized ASOs allows ASOs of thoroughly understood chemical classes to progress rapidly from *in vitro* testing to experimental use in humans after a single three-month good laboratory practice (GLP) toxicology study in a single species. The exposure of patients to experimental ASOs with such limited nonclinical data demands that each step in the process towards treatment be of the upmost quality. This begins with patient selection, based on complex risk/benefit assessment, through clinical exposure and evaluation of the performance of the ASO using the highest possible quality clinical outcome measures.

### Organs, chemical classes and routes of administration

Currently, n-Lorem accepts applications for patients who exhibit phenotypes resulting from a mutation in a gene that is expressed in the liver, kidney, central nervous system (CNS) or eye that is causal of the signs and symptoms currently displayed by the patient. With each of these organs, there is meaningful clinical experience and well-defined sub-organ pharmacokinetic/ pharmacodynamic (PK/PD) properties[15, 26–28]. Genetic targets in all four organs have been shown to respond to low doses of these drugs and the therapeutic margins are high, thereby enhancing the potential for safe and well-tolerated administration of experimental ASOs. For targets in the central nervous system, PS/PO 2′-MOEASOs are administered intrathecally (IT), approximately every 3 months. For targets in the liver PS 2′-MOE GalNAc ASOs are administered subcutaneously (SQ), approximately monthly. ASOs directed to the kidney are administered SQ approximately monthly. For targets in the eye, either PS 2′-MOE or PS 2′cEt are administered intravitreally (IVT) every 3 or 6 months.

### Quality processes

Research physicians can access an application for treatment on the n-Lorem website (www.nlorem.org) and submit on behalf of their patients.

Because we engage in n-of-1 treatment, each patient is assessed individually, and we consider three types of criteria, genotype, phenotype and administrative issues (Fig. 1). We determine if the pathogenic mutation is clearly understood and if other pathogenic mutations might contribute that they are clearly excluded as causal. Next, we determine if the mutation is amenable to ASO treatment. Typically, the most common mutation deemed unapproachable with ASOs is a null mutation. Frequently, we must perform additional experiments in our laboratory to be confident that the pathogenic mutation truly is well understood and amenable to ASOs. In parallel, the phenotype is assessed. We ask if the disease is manifested in one of the organs we currently treat, is the patient severely affected or likely to become so, what is the progression rate and is there near-term risk of death or loss of an organ. In addition, we determine if the cellular phenotype is well characterized and likely to be useful for assessing ASO treatment effects. Though we hope to be treating patients in countries other than the United States (U.S.), at present we accept only patients who can be treated in the U.S. if the prevalence of treatable patients is <30. Next, we determine if the treating physician and institution are willing and able to treat and evaluate the patient in a single patient clinical trial. Additionally, since we assess the effects of ASOs on the cellular phenotype, we need to be confident that we can access patient derived cells. Once we have assessed all the required information, we risk/benefit of treating the patient is considered and a detailed report of patient is presented to the Access to Treatment Committee (ATTC), a panel of individuals with expertise in diseases of the relevant organs, clinical care, and clinical trials of experimental agents. A typical ATTC presentation of a patient is shown in the supplemental materials. If the patient is deemed in urgent need of treatment, the application is prioritized as urgent. Otherwise, patients are treated on a “first come, first serve basis”. The ATTC then adds its perspective before a final decision is made about whether to develop an experimental ASO to treat this patient. Our goal is to present each patient to the ATTC within six weeks of receipt of the application. The median and mean time from receipt of application to the ATTC presentation are 56.3 days and 51.5 days, respectively (Fig. 2a). Importantly, we believe that the decision to treat can only be made in the context of each individual patient. Moreover, since ASO technology continues to advance, we think that this patient by patient approach is preferable to publishing guidelines that might result in denying a patient ASO treatment inappropriately.

**Figure 1.**
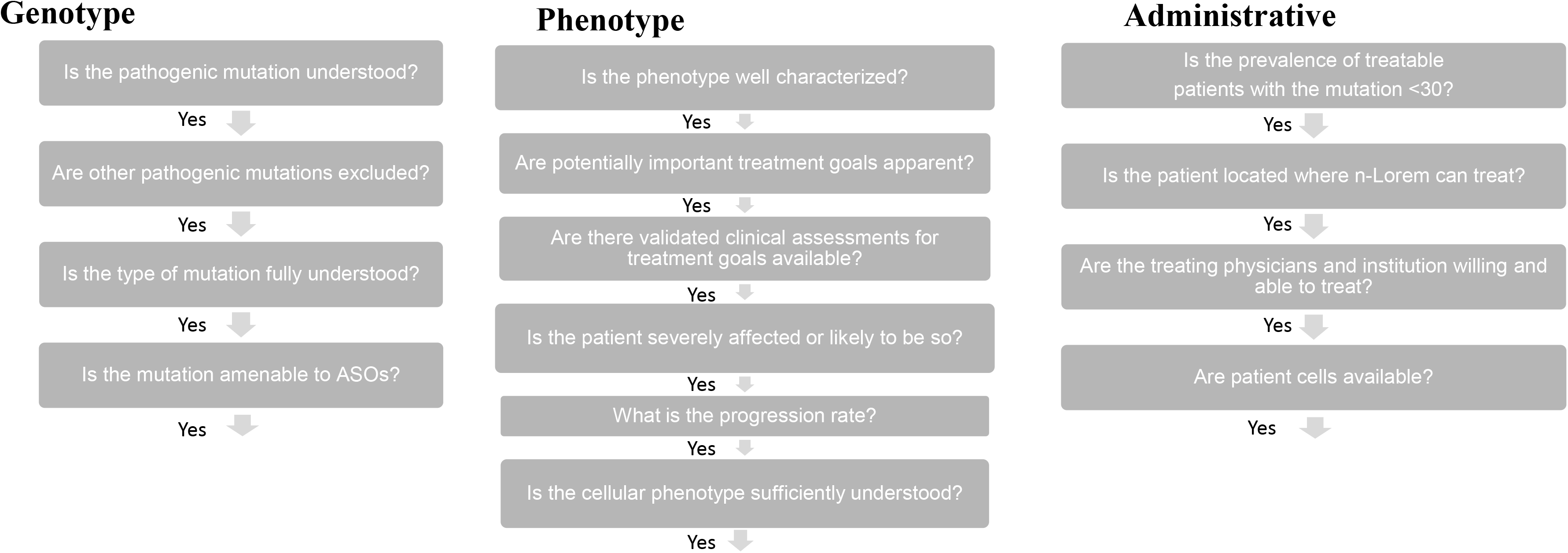
What patients should be treated with ASOs: Systematic personalized process. The n-Lorem team evaluates each application using the scheme shown. Then a formal presentation is prepared and presented to ATTC.

**Figure 2.**
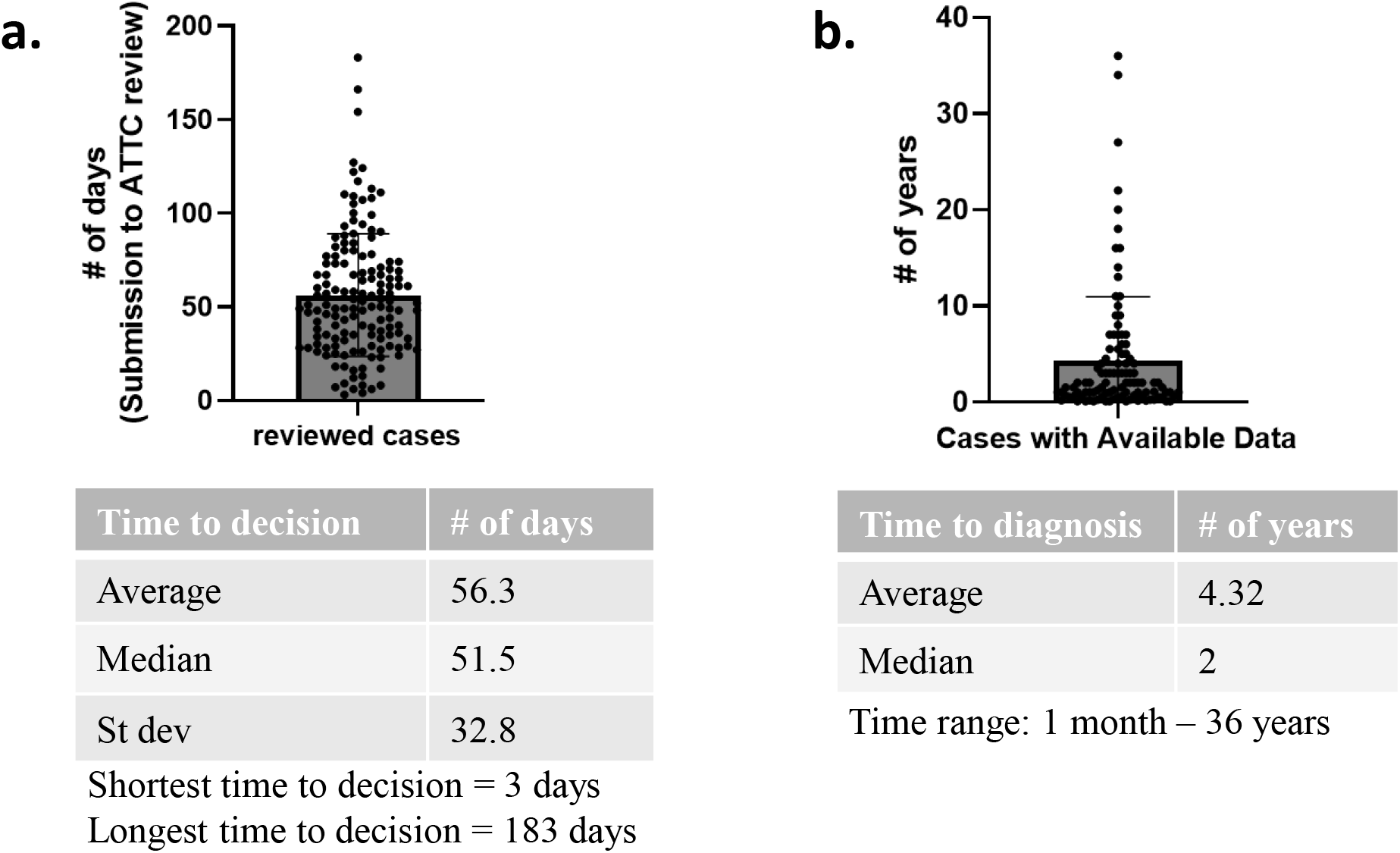
Times to diagnosis and decisions on applications for treatment. The bars on each graph represent the mean and the standard deviations of the means are shown in the figure. a. Time from submission of application to n-Lorem to treatment decision. b. Time from symptom onset to diagnosis

### Design, discovery and development of an optimal ASO

Given the limited nonclinical information available before initiating clinical treatment, it is essential that the individualized medicine derives from a thorough ASO design and screening process. Of course, screening processes differ depending on the organ involved and the degree to which molecular mechanisms resulting in ASO-induced phenotypes in the organ are understood. As experience is gained and mechanistic understanding advanced, screening approaches have evolved and become more efficient (Table 1). Today, the organ system that is least well understood is the CNS, meaning that screening for these intrathecally administered ASOs is the most cumbersome and requires that several hundred ASOs be evaluated *in vitro* and 20 or more be assessed in rodent tolerability studies. This table shows the typical number of ASOs screened at each step and the pass/fail criteria for each step. Obviously ASOs that advance from one step to the next must meet the pass/fail criteria. Typically, at the conclusion of *in vitro* screening, ∼20 PS ASOs are subjected to rodent tolerability screens. These screens vary depending on the organ to be treated but are similar to screening used for CNS ASOs.

Once an optimal ASO has been identified, it must be manufactured under good manufacturing practices (GMP), administered at least two dose levels for 13 weeks in the appropriate species, and then formulated in sterile vials. Each of these steps consumes time and involves multiple organizations in addition to n-Lorem. Our goal is to complete the entire process within 15-18 months (Supplemental Fig. 2). However, because the demand has substantially exceeded expectations, and despite our rapid growth, the median and mean times for acceptance of application to first dose are 23 months and 25 months, respectively. More than 50% of our patients present with heterozygous mutations in genes that encode proteins with essential functions, necessitating the design of allele-selective RNase H1 ASOs. Applications that require allele-selective ASOs for CNS diseases are often more challenging and may require more extensive screening and time.

### Assessment of ASO performance

Although clinical trials for single patient studies exist, these are typically cross-over designs in which a patient is treated for a period of time with a placebo or a reference agent, then with the experimental agent[29]. However, the long duration of action of modern ASOs, the severity of and the rate of progression of diseases in nano-rare patients obviate such designs. We, therefore, have created a new approach. Prior to initiating an effort to identify an optimal ASO for a particular patient, we work with the investigator and patient or parent to define primary, secondary and exploratory treatment goals. Validated clinical measures are selected to evaluate the status and rate of progression of symptoms in the patient during the time required to discover and develop the optimal ASO and thus provide each patient’s recent specific history. The treatment goals and clinical measures are reviewed and approved by a group of relevant experts, i.e. the Study Treatment Assessment and Review (STAR) committee. During treatment with the experimental ASO, the status and rate of progression of each agreed treatment goal are assessed and then compared to pre-treatment (recent patient-specific history) values. Obviously, as each study is, in effect, a first-in-human study, the primary objective of the trial is to assess safety and tolerability. Importantly, each study in a nano-rare patient has clear therapeutic goals as well, which are defined in the investigational new drug application (IND) for that ASO and patient. Adverse events are monitored by a Data Safety Monitoring Board (DSMB) and learnings from all individualized clinical studies will, over time, aid in further characterizing and improving personalized ASOs.

### Criteria to define a symptom as different from the symptom complex common to other patients with the equivalent mutation in the same gene

The diversity of mutations, genes and mutation-induced phenotypes makes defining a symptom as different from the phenotype common to a particular mutation in a specific gene complex, but important. In Table 3, we identify any symptom in a specific patient expressed in an organ other than the organ affected in shared phenotype as different, e.g., gastrointestinal (GI) symptoms in a patient who expresses a mutation with the primary phenotype being neurodegeneration. We also define as different from the shared mutationally caused phenotype any qualitatively different symptom in the organ that is affected in other patients who express the mutation, e.g., seizures that occur in a patient with a mutation that results in a common phenotype of progressive loss of muscle strength.

**Table 2:**
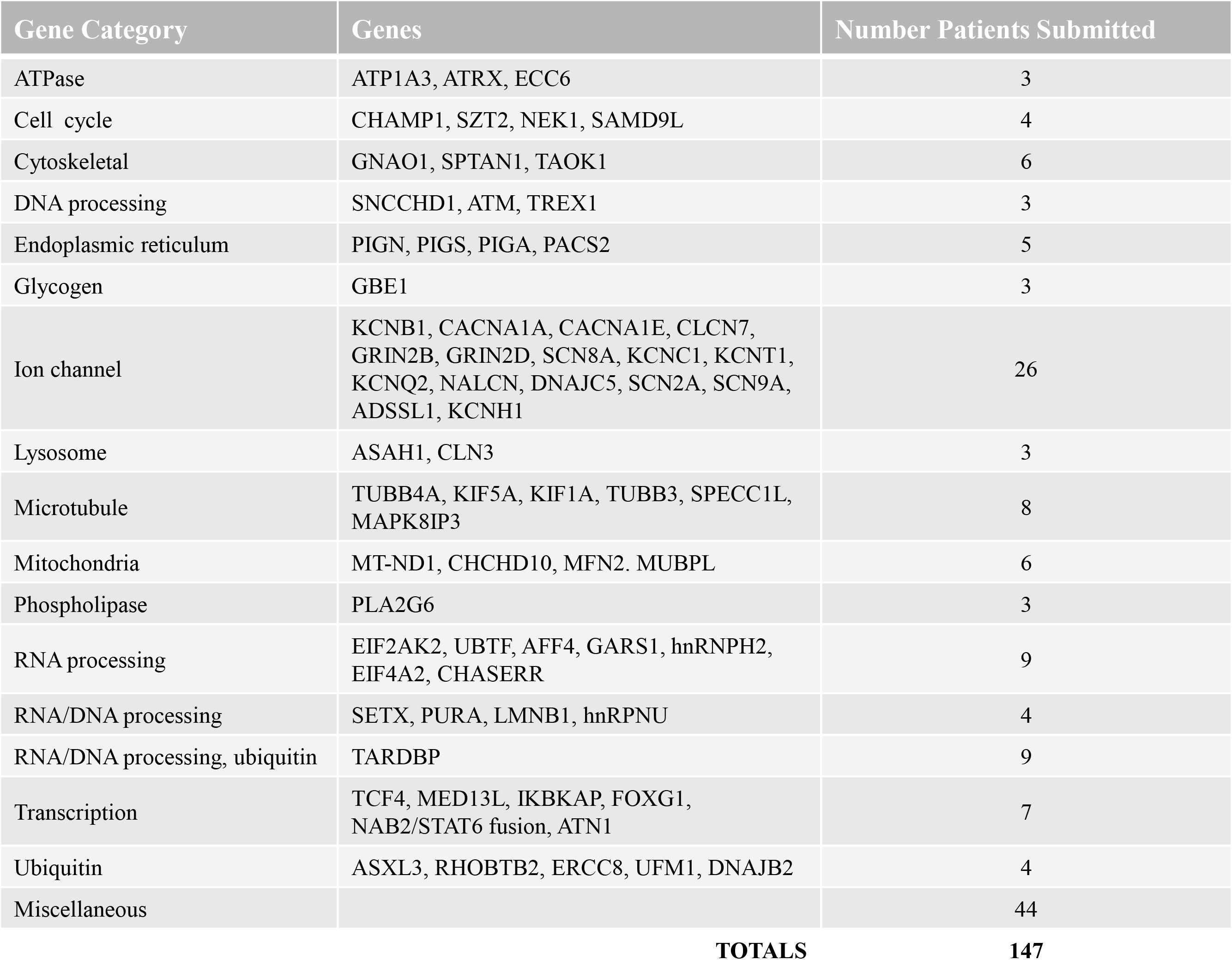
Functional Classification of Genes Submitted.

**Table 3:**
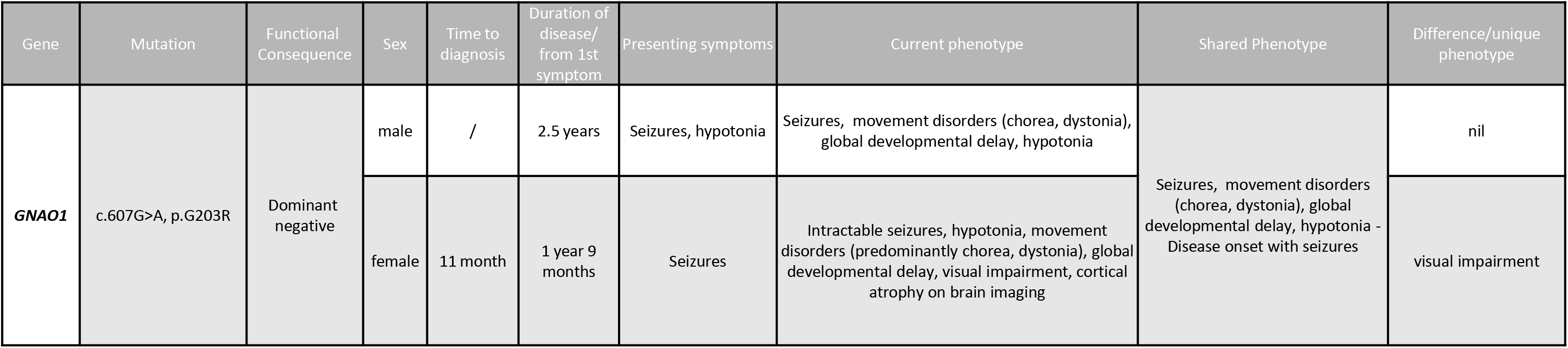
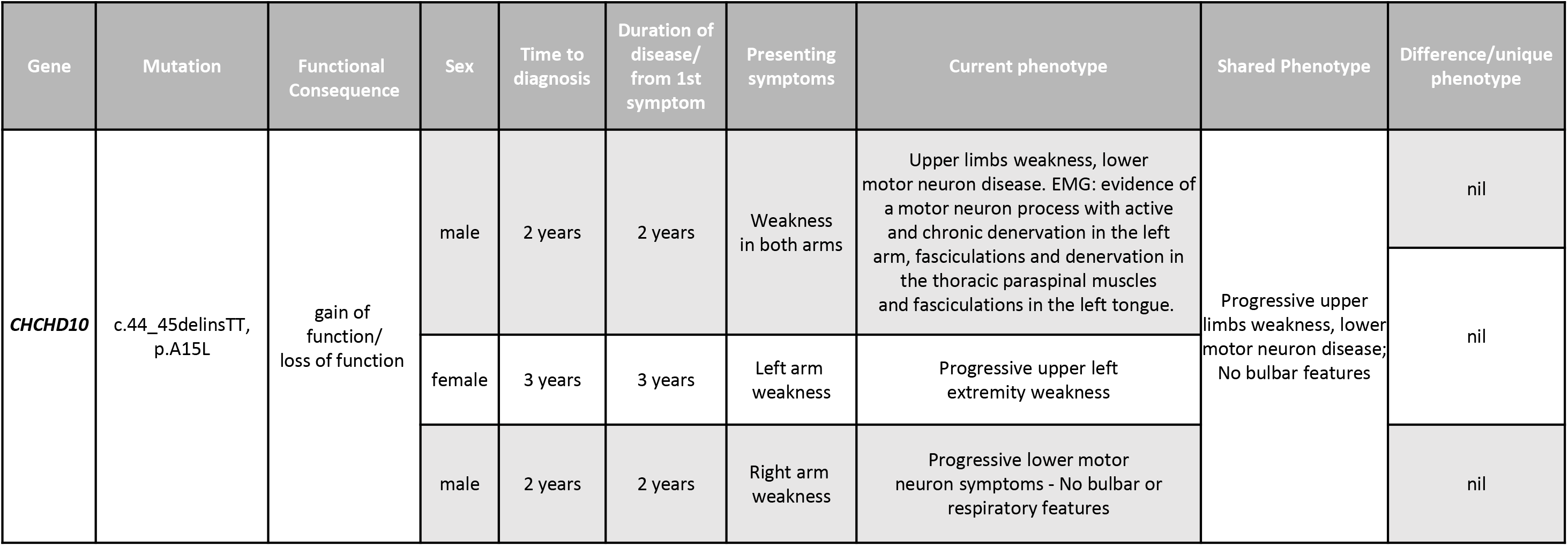
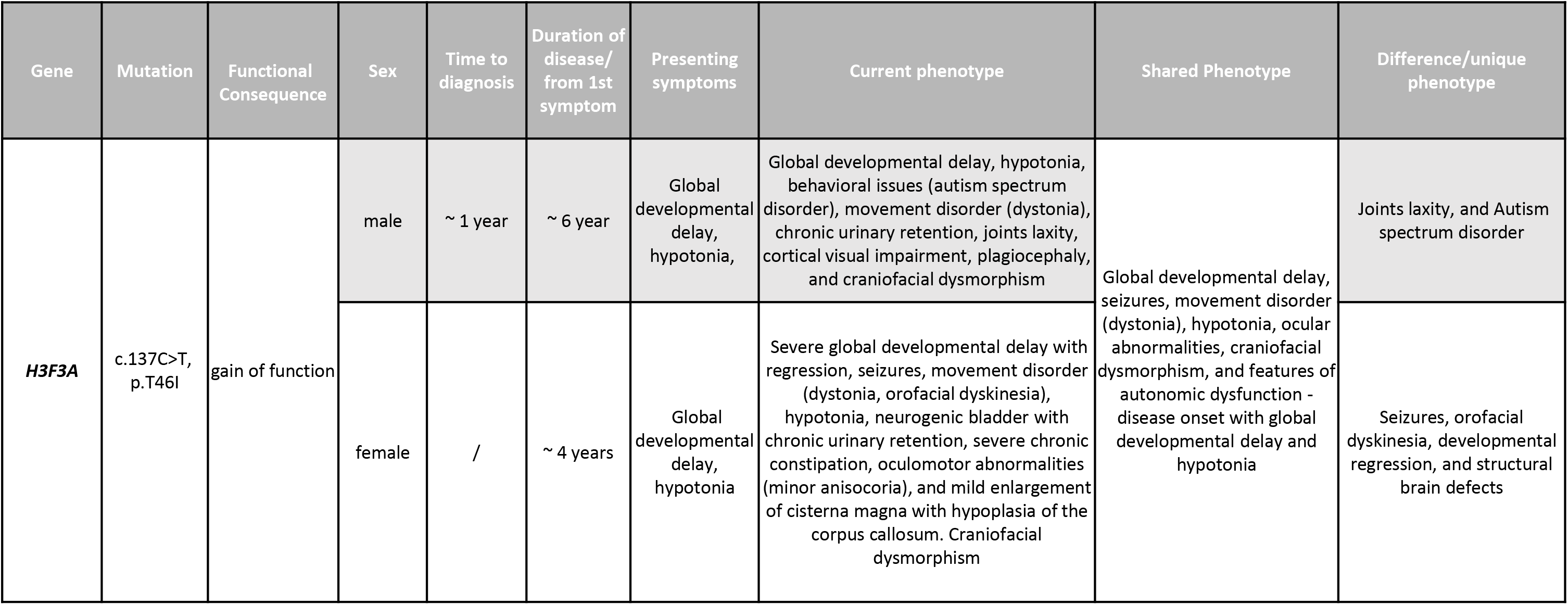
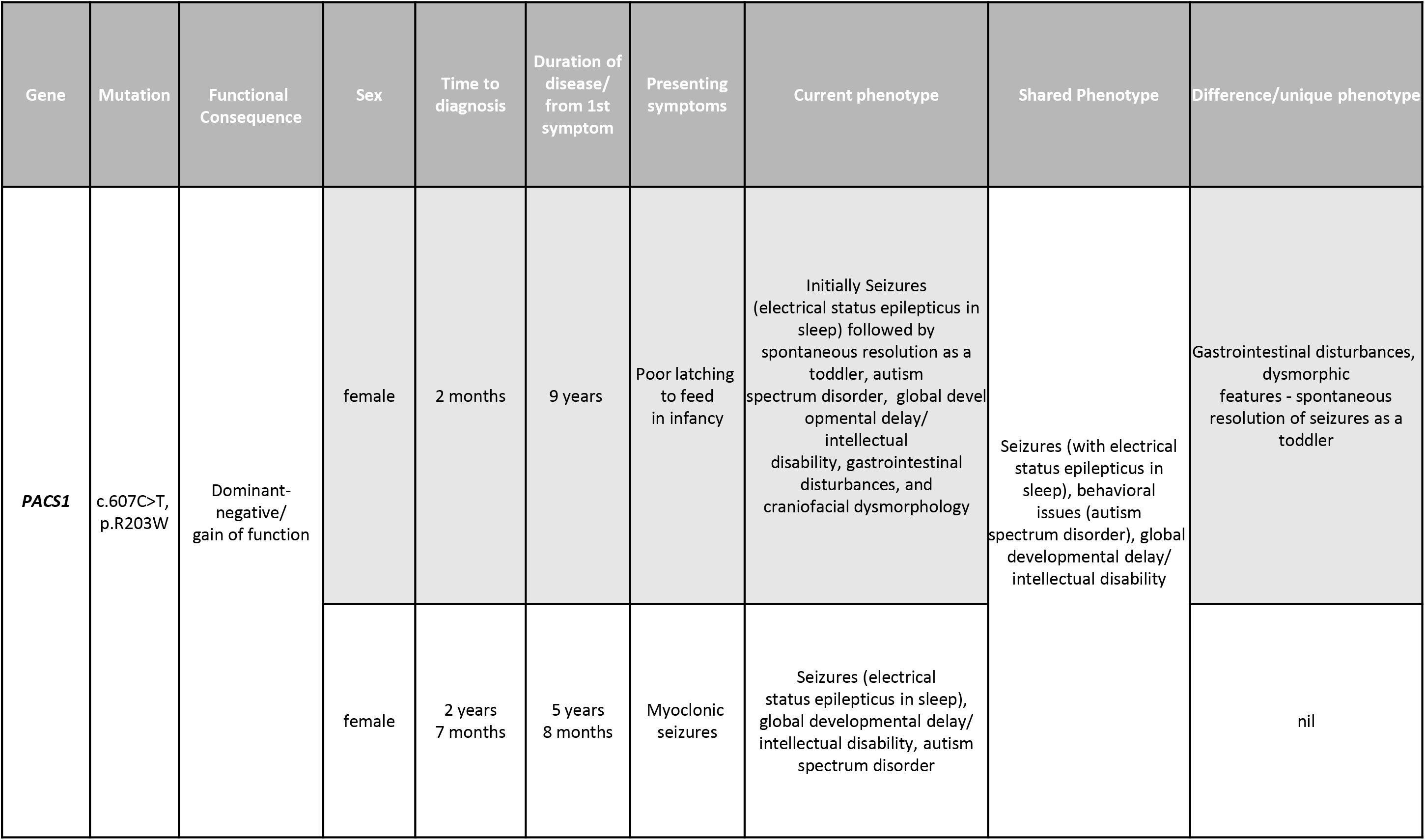
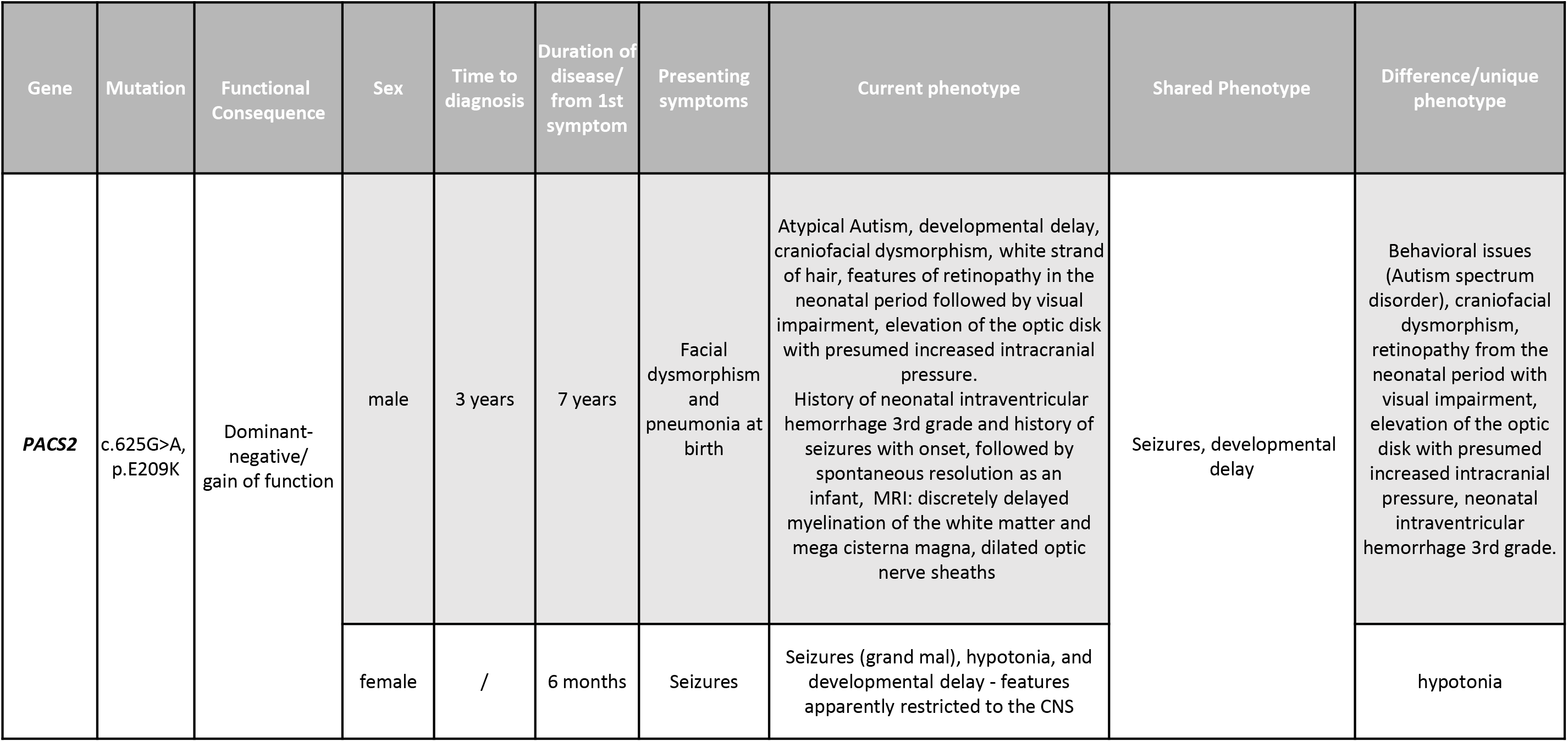
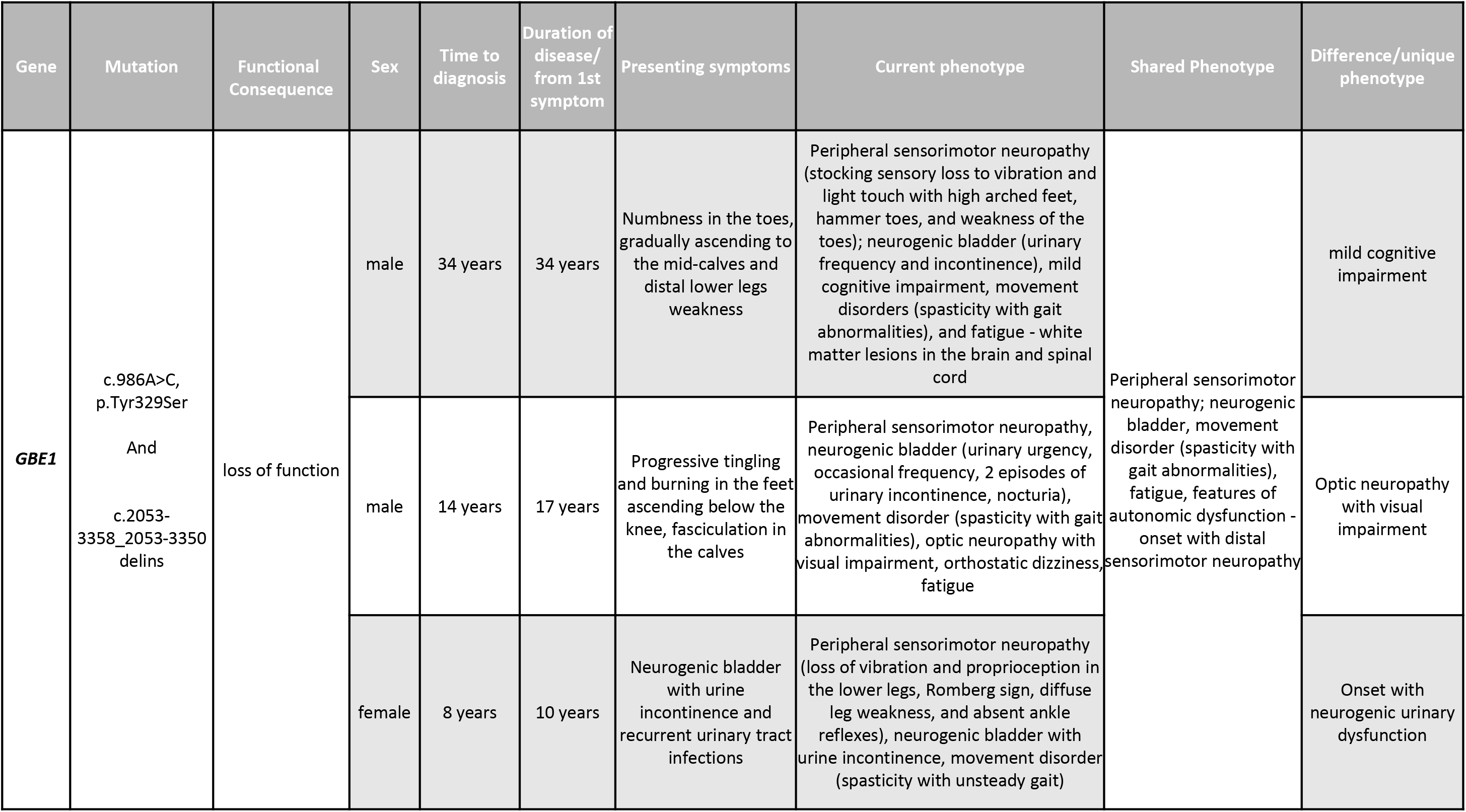
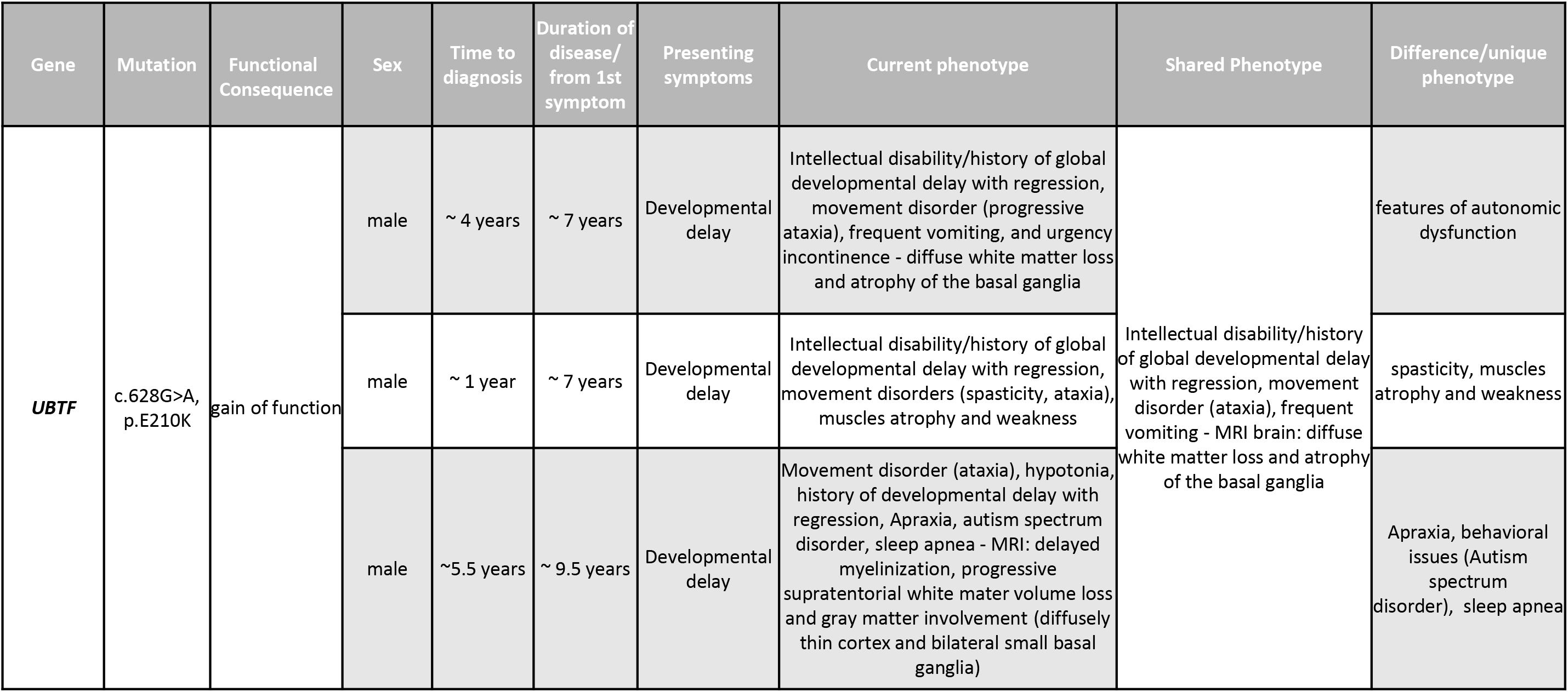
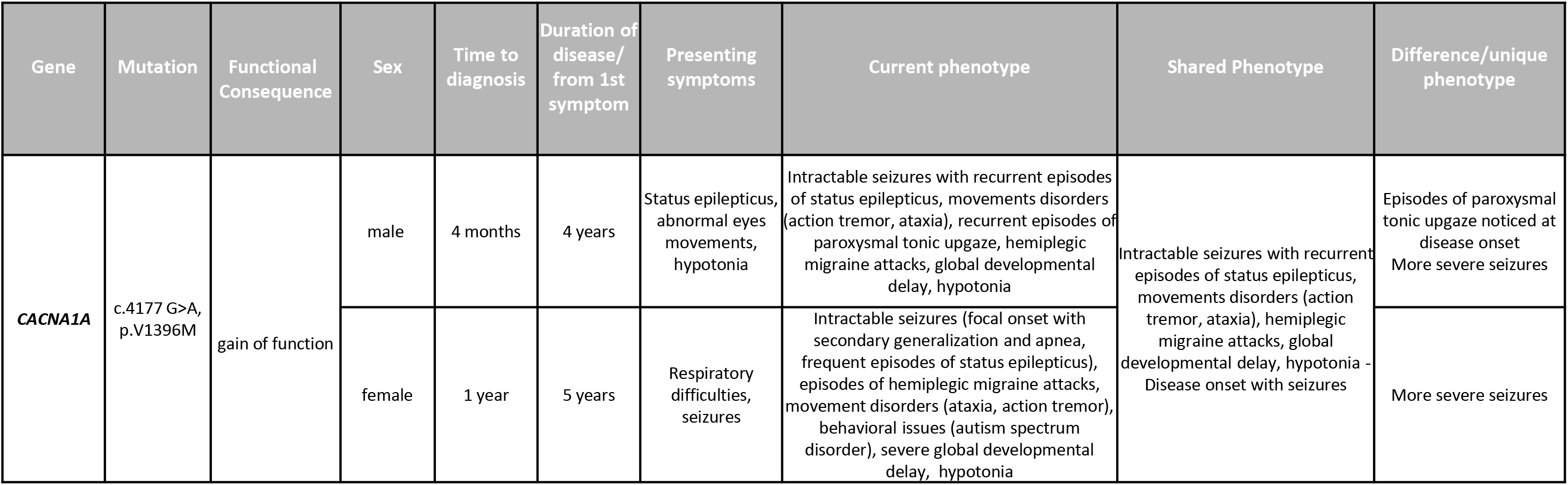
Genotype Phenotype Correlations. a. *GNAO1* patients with equivalent mutations. b. *CHCHD10* patients with equivalent mutations. c. *H3F3A* patients with equivalent mutations. d. *PACS1* patients with equivalent mutations. e. *PACS2* patients with equivalent mutations. f. *GBE1* patients with equivalent mutations. g. *UBTF* patients with equivalent mutations. h. *CACNA1A* patients with equivalent mutations.

## RESULTS

### Overall results

### Demographics

Of the 173 applications submitted as of January 2, 2023, 78 were accepted for potential treatment with an experimental ASO. Fourteen applications were on hold for a variety of reasons, including the need for more extensive genomic sequencing (Fig. 3a). Often, patients have only undergone whole exome sequencing and long read whole genome sequencing is often required for ASO drug discovery and development. Another common deficiency is a lack of definitive evidence about the nature of the mutation. For example, a patient who presented with a mutation in the *MAPK8IP3,* gene that encodes an adaptor protein involved in late endosomal movement along the microtubule network, has required more than a year of research to determine that the mutation is a toxic gain-of-function (TGoF) (manuscript in preparation). Another common challenge is to unequivocally demonstrate that the mutation posited to be the pathogenic mutation is indeed causative.

**Figure 3.**
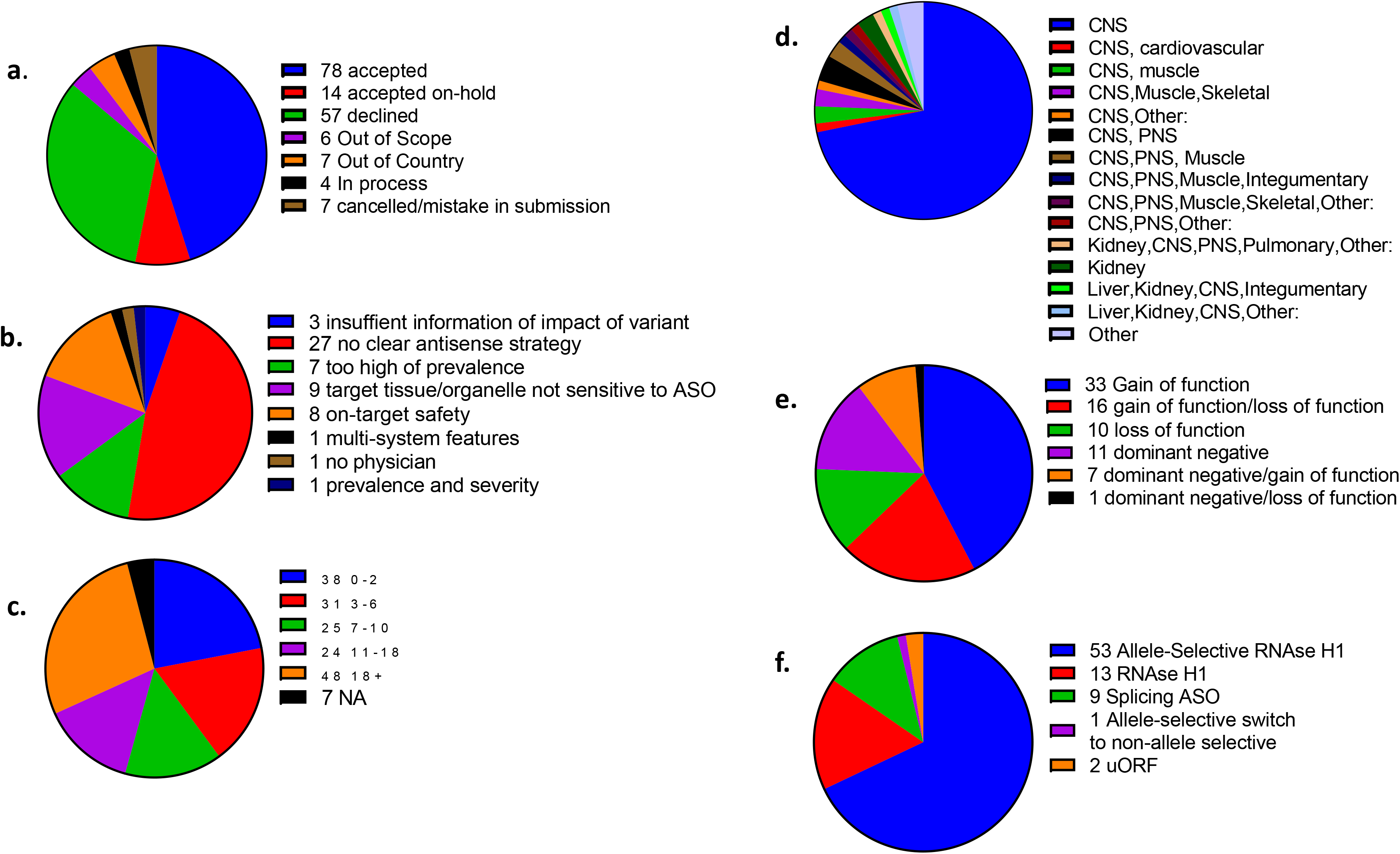
Characteristics of patients who submitted applications and antisense approaches for accepted patients. a. Total submissions and initial decision. b. Reasons for declined cases. c. Age distribution for submissions. d. Organs affected for accepted cases. e. Types of mutations for accepted cases. f. ASO strategy for accepted cases.

We are working with investigators to reduce the fraction of incomplete applications. Fifty-seven applications were declined, with the most common cause being a lack of an antisense approach to the mutation (Fig. 3b). The ages of patients ranged from infants to >65 years of age. Importantly, 48 patients who applied for treatment are >18 years old and the age distribution of applications accepted for potential ASO treatment was similar (Fig. 3c). That many of these patients had mutations with long latencies is an important observation discussed in a later section. The organs affected were heavily weighted to the CNS for both total applications and accepted cases (Fig. 3d). Obviously, nano-rare mutations are expressed in the other organs, so the preponderance of CNS applications simply reflects a lag in engaging those communities of physicians and patients. For applications accepted, the post-RNA binding mechanisms of action (Fig. 3e) for which ASOs were designed (Fig. 3f) reflected the nature of the mutations and the available options in ASO strategies. Approximately 68% of patients required an allele-selective RNase H1 ASO.

As shown in Supplemental Figure 2, our goal is to initiate dosing within 18 months of acceptance of the application. To date, that goal has not been achieved. The median and mean time from acceptance to initial dose are 23 and 25 months, but these are not meaningful statistics as they represent the first 8 INDs approved by the FDA. That said, two factors contributed to the delays; first, the total number of cases accepted has greatly exceeded our expectations, requiring substantial growth in staff and facilities to meet the demand. Second, because many patients have heterozygous mutations in essential genes, a large number of allele-selective PS 2′-MOE ASOs were needed, a process which is more time-consuming than discovering non-allele selective ASOs.

### Diagnostic journey

The patients reported here present the first opportunity to directly assess the time from symptom onset to diagnosis for the fraction of nano-rare patients who actually achieve a diagnosis. We have sufficient data to determine the time required to achieve a diagnosis for 102 patients. The range in time was 1 month to 36 years. The mean and median times from symptom onset to diagnosis were 4.32 years and 2 years (Fig.2b). This reflects the enormous challenges that the rarity of these mutations imposes on taking the obligatory first step toward treatment, achieving a genetic diagnosis, the identification of the mutation responsible for the patient’s phenotype. The mean and median times to diagnosis are of less value than the enormous range in times to diagnosis in demonstrating how idiosyncratic the diagnostic journeys are for nano-rare patients.

### Genotypes/phenotypes

In Table 2, we present the classes of genes with nano-rare mutations represented in the 173 patients who applied for treatment. Cases that had multiple genes associated with the same process or pathway are listed. Forty-four cases are so unique that no other cases were found to be able to group them. A database that couples detailed genomic and phenotypic data on individual patients that can be identified will facilitate long term follow-up to better assess how genotypically dependent phenotypes vary over time. However, our database has significant limitations. First, the number of evaluable patients is still small. Second, applications are heavily weighted toward genes with known functions in the CNS, and we cannot exclude selection bias based on perceptions about the limitations of antisense technology. As the database expands, itsvalue will increase.

Diverse genes and functional families are represented. Given the preponderance of CNS diseases, it is not surprising that mutations in many ion channel genes are significantly represented. Also of note are mutations that affect endosomal trafficking along microtubules in axons. Not surprisingly, multiple steps in neurotransmitter function are represented. Multiple nucleic acid binding and chaperone proteins, transcription factors and proteins involved in endoplasmic reticulum and Golgi functions are also represented. The large number of patients with mutations in *TARDBP* probably represents selection bias introduced by a very active patient advocacy group. We compiled detailed genotypic information and molecular causes for mutations for which we had more than one patient that applied for treatment and for whom we have sufficient phenotypic data to support a detailed analysis (Supplemental Table 1).

As expected, the organs affected and phenotypes vary widely for different mutations in the same gene, often resulting in divergent phenotypes, and the age at symptom onset varies. Though not surprising, this information is of interest and the value of these comparisons will increase as new patients are evaluated and followed. On the other hand, mutations in genes with similar functions such as ion channels, or genes involved in endosomal migration along microtubule networks often appear to result in similar phenotypes (data not shown).

### Phenotypes associated with the same mutation in the same gene can vary

While it is well known that different mutations in the same gene can result in widely divergent phenotypes, in Table 3, we asked how phenotypes vary in patients with the same mutation in the same gene. For example, (Table 3a) the phenotypes of two patients with the same mutation in the guanine nucleotide binding alpha protein O1 (*GNAO1*-c.607G>A, p.G203R)[29, 30], displayed a similar phenotype except that one patient experienced some severe vision issues while the other did not. Given that the patients are similar in age, it is unlikely that age would have contributed to these differences. The phenotypes of three patients with the same mutation in the coiled-coil-helix-coiled-coil-helix domain-containing protein 10 (*CHCHD10*), a gene that encodes a protein required for normal mitochondrial function[31] and associated with a range of syndromes including Parkinson’s disease, (PD), Alzheimer’s disease (AD), cerebellar ataxia, amyotrophic lateral sclerosis (ALS) or frontal temporal dementia (FTD)[32], are compared in Table 3b. All three patients had equivalent pathogenic variants that resulted in mixed gain and loss of function characteristics but, no obvious changes in the basic functional properties of the protein. All three patients presented with upper limb weakness between the ages 46 and 63 years. Aside from the variation in onset of symptoms and slight variances in muscle groups affected, the syndromes were quite similar.

Arguably more impressive and interesting is the diversity of phenotypes associated with mutations in Histone variant 3F3 (*H3F3.3A or B*) (Table 3c). Though the coding sequences of *H3F3A* and *B* are identical, they have unique regulatory components that result in differential expression in various tissues. H3.3 marks actively transcribed genes and somatic mutations have been known for some time to cause a variety of CNS malignancies. However, germ-line mutations in these genes are associated with progressive neurological dysfunction and, to date, no malignancies have been reported[33]. The mutations associated with neurodegeneration are missense in the coding region and thought to locally derange critical post-translational modifications. However, the range of phenotypes observed in the 46 patients identified with these mutations is enormous. The core phenotype consists of global developmental delay, hypomyelination, callosal anomalies, cortical atrophy and craniofacial abnormalities, but not all patients express these traits. Other manifestations include a variety of motor dysfunctions, oculomotor abnormalities and congenital defects in many organs. Though our two patients express equivalent mutations and had many overlapping features, they also differed, with one individual experiencing a more severe phenotype with seizures, developmental regression, movement disorders, and structural brain anomalies in addition to core features[33].

Similarly, the two patients with identical phosphofurin acidic cluster sorting protein 1 (*PACS1*) mutations shared common features, but one patient displayed GI dysfunctions, dysmorphic features and seizures which resolved when they were toddlers (Table 3d). The two patients with identical phosphofurin acidic cluster sorting protein 2 (*PACS2*) mutations displayed syndromes that differed significantly (Table 3e). Three patients with equivalent complex mutations in the 1,4- alpha-glucan branching enzyme 1 (*GBE1*) gene also showed meaningful phenotypic differences (Table 3f) as did equivalent mutations in the upstream binding transcription factor (*UBTF*) gene (Table 3g). A final example (Table 3h) is calcium channel, voltage-dependent, P/Q type, alpha-1A subunit (*CACNA1A*), a gene that encodes a voltage dependent calcium channel expressed primarily in the brain. Mutations in this gene result in severe disorders that include severe epilepsy and movement disorders. Two patients of approximately the same age with the same toxic gain of function mutation share many elements of the expected phenotype, but one patient expressed significantly more problems with movement, while the other had more severe epilepsy.

### Different, but functionally equivalent, mutations can result in similar phenotypes

Our data also present the opportunity to more thoroughly evaluate whether different mutations with shared molecular pathological effects, e.g., TGoF, in the same gene result in similar phenotypes (Table 4). The best example of this are the missense mutations in the gene that encodes the TAR DNA Binding Protein 43 (*TARDBP* gene and TDP43 protein). A number of these patients display similar clinical features, but a few patients present with slight variations in phenotype.

**Table 4:**
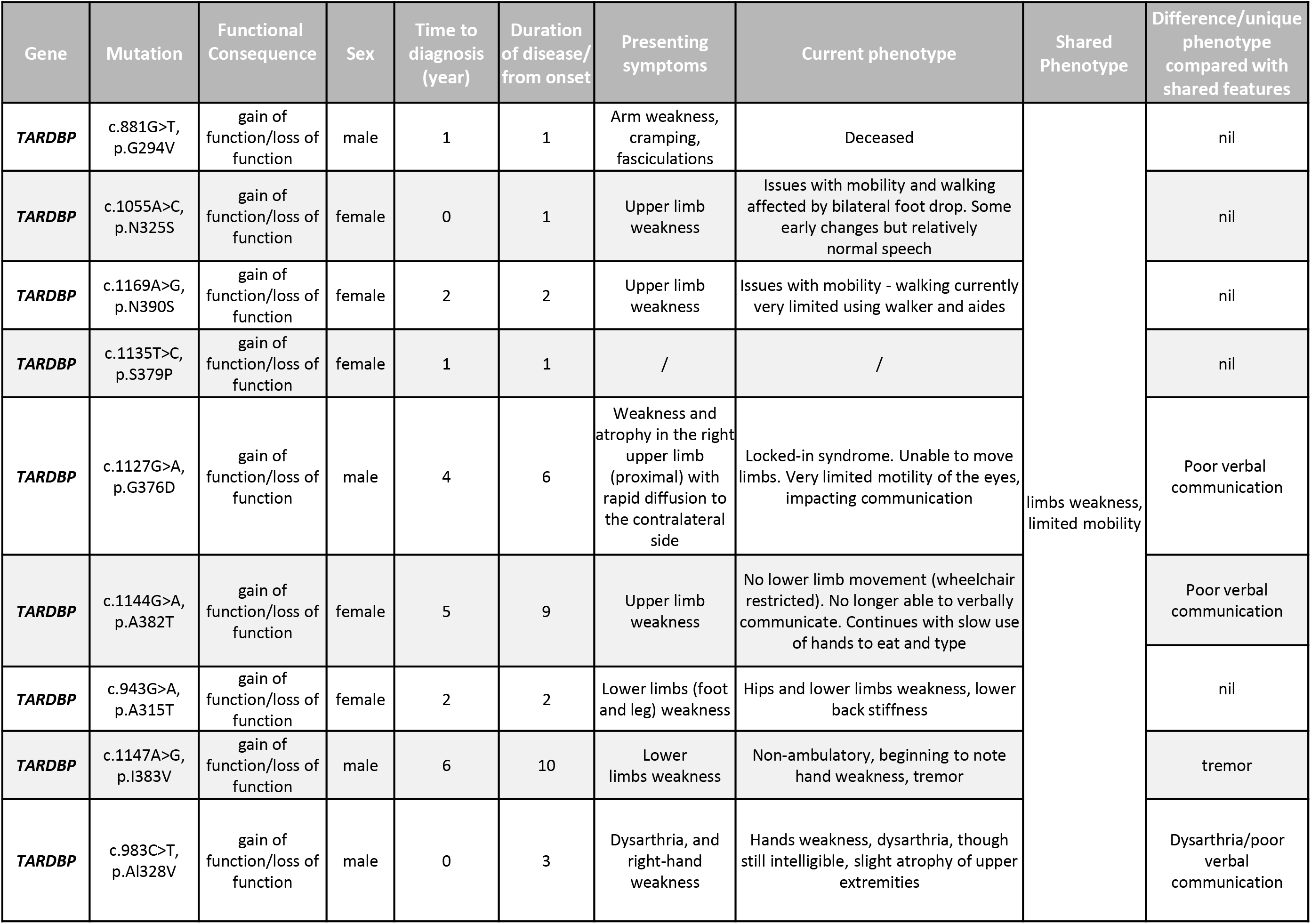
Comparison of the Phenotypes of Patients with Different Mutations in the Same Gene. *TARDBP* different, but functionally equivalent mutations.

TDP43 is a particularly instructive example of a protein whose effects may be driven by multiple roles due to specific domains that serve different functions, including interactions with other proteins. The many domains that constitute TDP43 include a nuclear localization and nuclear export domain, two RNA binding domains and c-terminal domains that are essential for homo- dimer formation and the potential to form toxic aggregates[34] and most mutations are localized to the c-terminal domain. These result in a loss of function in nuclear RNA metabolic processes and a toxic gain of function in the cytoplasm associated with toxic aggregates in stress granules and other sites. Given the pleiotropic roles of the protein and the multiple pathogenic effects of toxic aggregates in the cytoplasm, one would expect mutations to cause diverse phenotypes and be involved in multiple diseases and that is exactly what has been observed in the literature and in our patients. However, to a large extent, different, but functionally equivalent mutations in the same gene result in different phenotypes (Table 5, Supplemental Table 1).

**Table 5:**
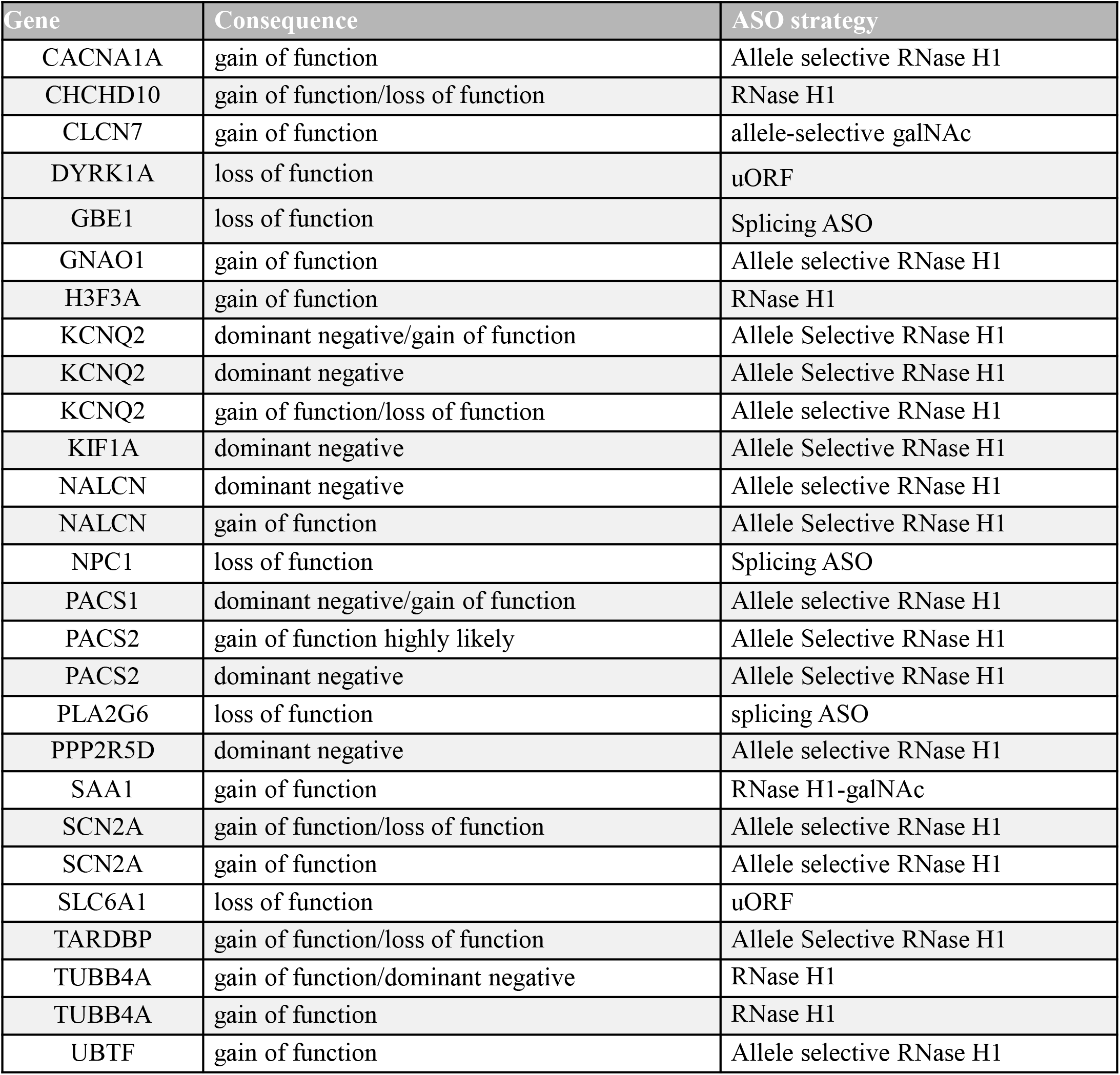
Genes. Types of Mutations and Antisense Strategies for Accepted Patients.

| Gene | Consequence | ASO strategy |
| --- | --- | --- |
| CACNA1A | gain of function | Allele selective RNase H1 |
| CHCHD10 | gain of function/loss of function | RNase H1 |
| CLCN7 | gain of function | allele-selective galNAc |
| DYRK1A | loss of function | uORF |
| GBE1 | loss of function | Splicing ASO |
| GNAO1 | gain of function | Allele selective RNase H1 |
| H3F3A | gain of function | RNase H1 |
| KCNQ2 | dominant negative/gain of function | Allele Selective RNase H1 |
| KCNQ2 | dominant negative | Allele Selective RNase H1 |
| KCNQ2 | gain of function/loss of function | Allele selective RNase H1 |
| KIF1A | dominant negative | Allele Selective RNase H1 |
| NALCN | dominant negative | Allele selective RNase H1 |
| NALCN | gain of function | Allele Selective RNase H1 |
| NPC1 | loss of function | Splicing ASO |
| PACS1 | dominant negative/gain of function | Allele selective RNase H1 |
| PACS2 | gain of function highly likely | Allele Selective RNase H1 |
| PACS2 | dominant negative | Allele Selective RNase H1 |
| PLA2G6 | loss of function | splicing ASO |
| PPP2R5D | dominant negative | Allele selective RNase H1 |
| SAA1 | gain of function | RNase H1-galNAc |
| SCN2A | gain of function/loss of function | Allele selective RNase H1 |
| SCN2A | gain of function | Allele selective RNase H1 |
| SLC6A1 | loss of function | uORF |
| TARDBP | gain of function/loss of function | Allele Selective RNase H1 |
| TUBB4A | gain of function/dominant negative | RNase H1 |
| TUBB4A | gain of function | RNase H1 |
| UBTF | gain of function | Allele selective RNase H1 |

In Supplemental Table 2, we list the genotypes and phenotypes for all patients with sufficient information. To our knowledge, this is the first compilation of genotypes coupled to phenotypes of patients with extremely rare mutations that also can identify specific patients. This table is meant to serve as a resource should other patients with the same or different mutations in any of these genes be identified. Importantly, for any mutation and gene accepted, we may have an ASO that targets that mutation, or a common SNP, which could be beneficial to others with the same mutation. We plan to publish this information and update it regularly.

### ASO mechanisms vary depending on the nature of the mutation

The appropriate post-RNA binding mechanism for each patient is summarized in Table 5. The assessment of the feasibility of ASO treatment of a mutation, the whether the target tissue is appropriate, and the prevalence of the pathogenic mutation and the risk/benefit of treatment are all considered in the decision to accept or reject an application for treatment. The integration of the various types of analyses enables an informed decision about whether to treat a patient and what ASO strategy is appropriate.

Full characterization of the mutation, proof that the mutation is truly causative, assessment of the proximal molecular pathological event, detailed phenotypic assessment, prevalence of mutation, organ affected, severity of disease, and rate of progression are considered to determine the benefit/risk of experimental ASO treatment of each patient. All the information available and the recommendation to treat or not are presented to and discussed with the ATTC. We believe that this detailed process is essential to maximize benefit and minimize risk.

### Exemplary patients

To provide more tangible information to better define the patient-by-patient process that leads to ASO treatment and early patient-specific clinical data, we present three exemplary patients: two patients with primary disease manifestations in the CNS who require allele-selective ASOs, and one with renal amyloidosis who is being treated with a non-allele-selective RNase H1 ASO. Though we have several ASOs that alter RNA splicing in development, we do not yet have sufficient clinical exposure to assess benefit. These individuals also demonstrate the complexity of the genetics and clinical syndromes of nano-rare patients. Using current methods and ASO designs, the design and identification of allele-selective RNase H1 active ASOs is more challenging because the ASO must be potent, allele-selective and well tolerated. To design an allele-selective ASO, one must identify non-pathogenic SNPs and screen ASOs around each SNP to select ASOs that selectively reduce the pathogenic mutant RNA and protein (for review see, 14- 18).

### *KIF1A* Patient

A female *KIF1A p*atient who has a *de novo* heterozygous c.914C>T variant in kinesin 1A (*KIF1A*). *KIF1A,* which is expressed primarily in the CNS, produces an essential microtubule motor protein that is responsible for anterograde movement of cargo-laden vesicles in neurons [35]. While most pathogenic variants occur in the motor domain, pathogenic variants are rarely are reported in other domains. Variants in *KIF1A* can cause a spectrum of neurological phenotypes that include movement disorders, epilepsy, painful neuropathy, developmental delays and intellectual disability [35, 36]. The patient demonstrated global developmental delays, progressive spasticity and peripheral neuropathy, and seizures. The patient’s spasticity contributed to many falls prior to treatment, and progressed from being largely ambulatory to requiring a wheelchair most of the time. The patient had difficulty with expressive language and speech fluency and articulation — frequently displayed irritability.

The patient has been treated with an allele-selective RNase H1 ASO, PS/PO 2′-MOE “gapmer” ASO (Table 6) designed to a non-pathogenic polymorphism in *KIF1A* (Fig. 3a) that was evaluated *in vitro* using patient iPSc-derived neurons. The ASO that was selected exhibited a good potency, safety and tolerability profile, and displayed a high degree of allele selectivity (Fig. 3b). Nonclinical development study results are presented in Supplemental materials (Supplemental Material – Detailed methods and results for exemplary patients) [37, 38].

**Table 6:** ASOs Used to Treat the Exemplary Patients. Color code for ASO chemical modifications: black = unmodified deoxyribose (2 H; DNA); orange = 2 methoxyethyl (MOE). Unmarked backbone linkages = phosphorothioate (PS); linkages marked with o = normal phosphodiester (PO). mC = 5-methylcytosine

| ASO | Sequence and chemistry | Target |
| --- | --- | --- |
| nL-KIF1-001 | AToGoToAomCATTTTmCTTTGoToTGmC | KIF1A |
| nL-SCN2-001 | TGomComCoAoAmCAATGTAmCAAoGoGGT | SCN2A |
| nL-SAA1-001 | GalNAcoAmCmCmCATTGTGTAmCmCmCTmCTTmC<br>mC | SAA1 |

Treatment was initiated at 20 mg IT and the dose-escalated gradually up to a dose of 100 mg. The patient has been treated for ∼15 months with no ASO-related serious adverse events, ASO-related adverse event, nor any emergent changes in the composition of the cerebrospinal fluid or other standard laboratory tests. The primary treatment goal to reduce the frequency and severity of seizures assessed and reported by caregivers in a seizure diary. The frequency and severity of seizures declined significantly beginning after the second dose and has been maintained throughout the treatment period (Fig 4a). Further, it was reported that the patient is more alert and present with treatment, and this is reflected by an overall improvement in the patient’s quality of life [39]. Of note, the specific domains of change demonstrate the dimensions of improvement. For example the patient is better able to express needs, shows more enjoyment in activities, has more energy, is in a better mood, shows less signs of anxiety, and shows less aggression since being treated with the ASO (Supplemental Table 4). While mobility only shows minor improvement on the clinically assessed six-minute walk test — rapidly recovered the ability to walk and demonstrated a dramatic reduction in falls even though the patient was much more mobile and ambulatory [39]. Additionally, reported by caregivers that there has been a significant decrease in pain—appearing to wear off near the end of the time between doses. Lastly, after ∼12 months of treatment, caregivers were asked to rate the severity of the patient at study start, and after one year of treatment, and to rate the improvement of the patient (see Supplemental Material for additional details). Both rated the patient as having ‘much improved’ and being ‘moderately ill’ after one year of treatment as compared to ‘severely ill’ or ‘markedly ill’ at study initiation (Supplemental Table 5).

**Figure 4.**
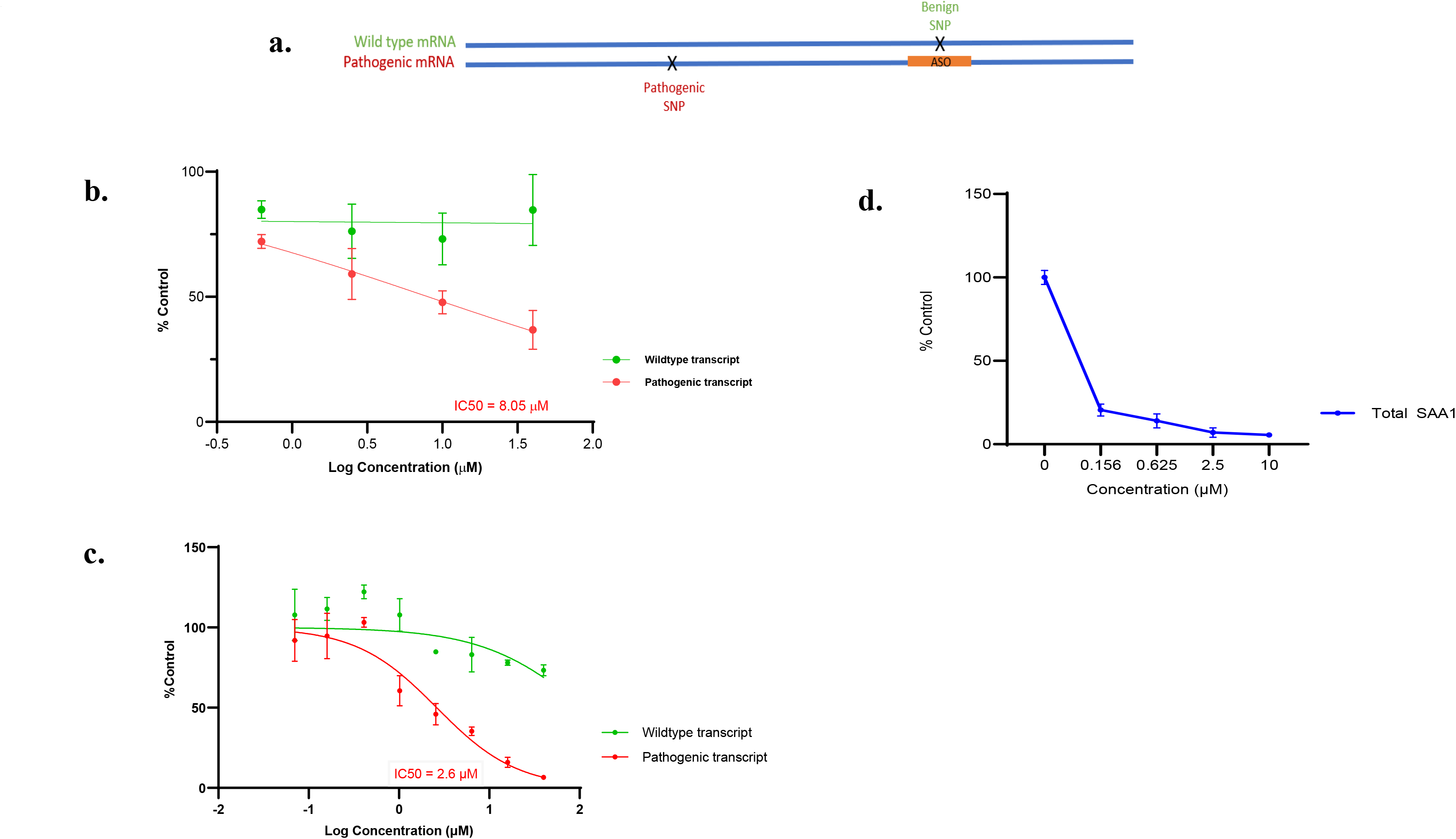
ASO design and dose-response curves for ASOs for exemplary patients. a. Schematic representation of an allele selective ASO designed to a SNP trans of the pathogenic mutation. b. Dose response analysis of nL-KIF1-001 in patient ipsc-derived neurons. c. Dose response analysis of nL-SCN2-001 in patient ipsc-derived neurons. d. Dose response analysis of nL-SAA1-001 in A431 cells.

**Figure 5.**
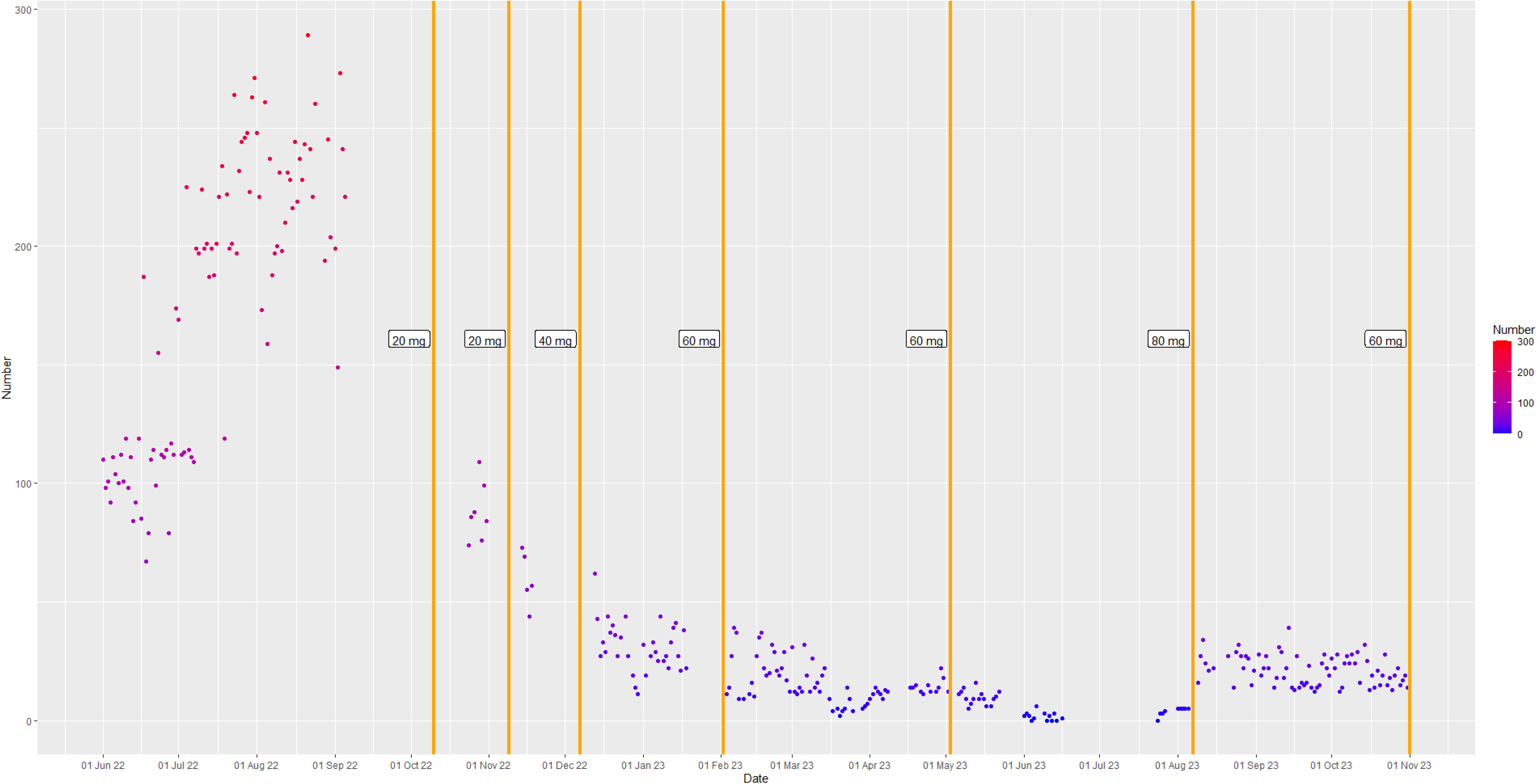
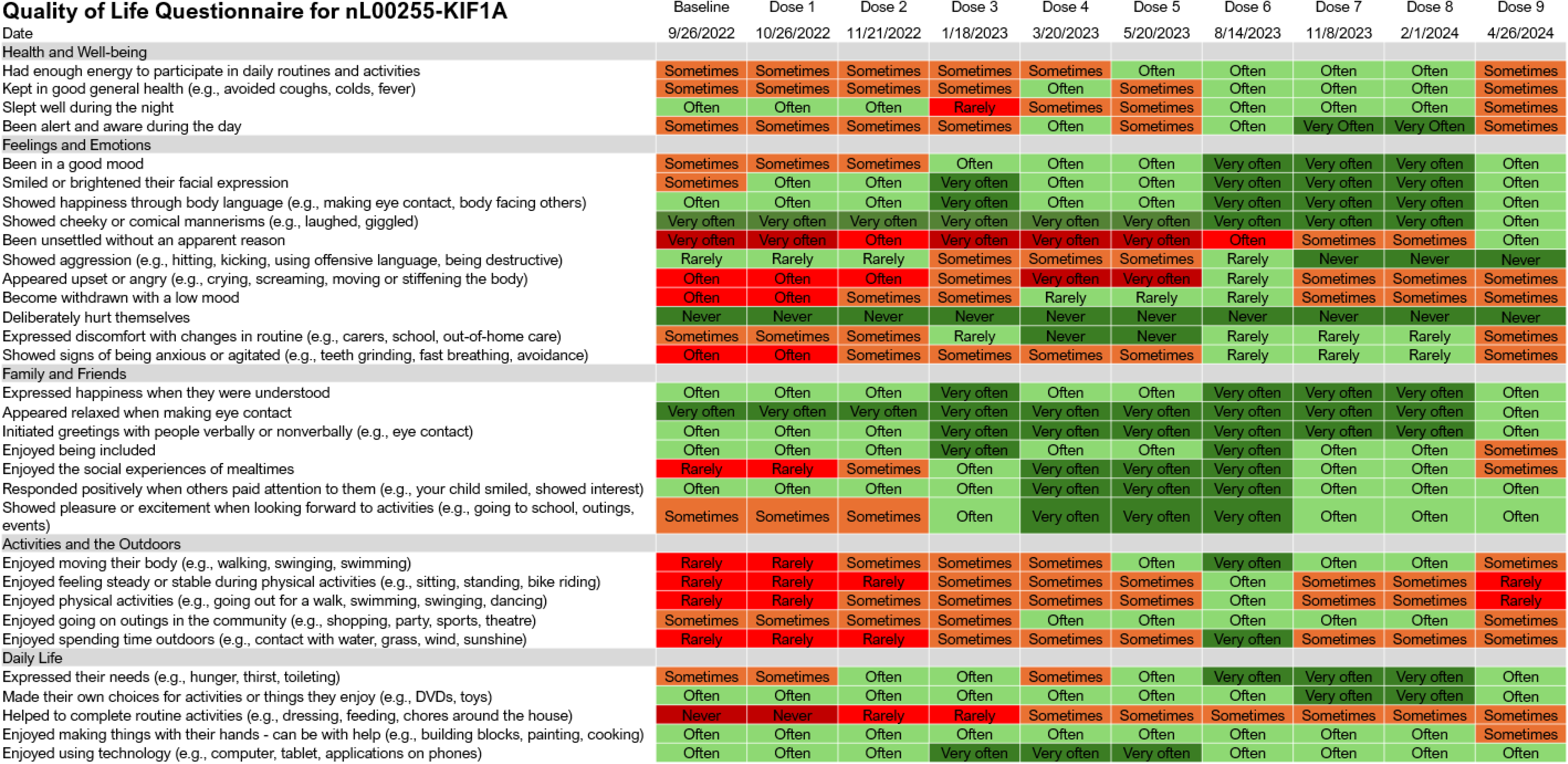
Seizure frequency over time post-ASO in two patients with previously intractable seizures. a. Seizure count over time in the KIFA patient . b. Seizure count over time in the SCN2A patient . Dose adjustments of rescue seizure medications are noted in green; DS = dose skip, DR = dose reduction, PM (A) – afternoon, PM –(C) - evening, AM – morning; Rescue medications noted in orange include phenytoin (P) or clonazepam (C). Phenytoin levels in blue are given in micrograms per mililiter.

**Figure 6.**
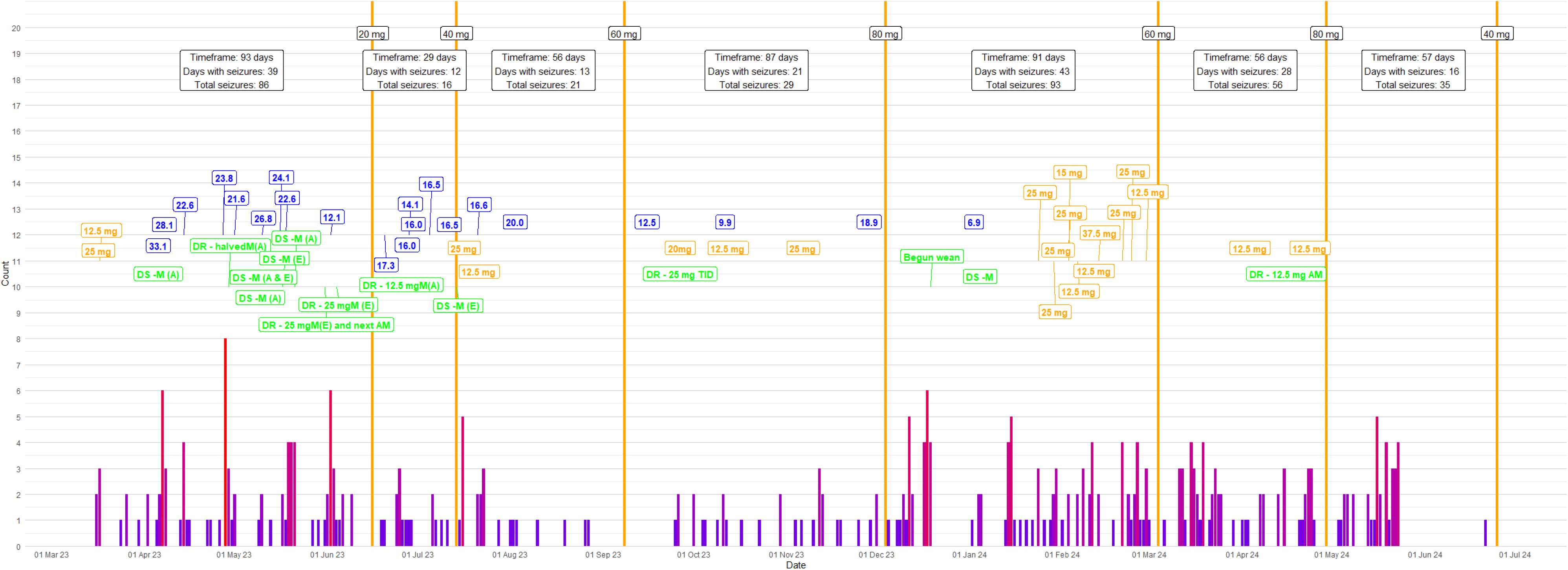
Seizure count overtime in the SCN2A patient. Dose adjustments of rescue seizure medications are noted in green; DS = dose skip, DR = dose reduction, PM (A) – afternoon, PM – (C) - evening, AM – morning; Rescue medications noted in orange include phenytoin (P) or clonazepam (C). Phenytoin levels in blue are given in micrograms per milliliter.

**Figure 7.**
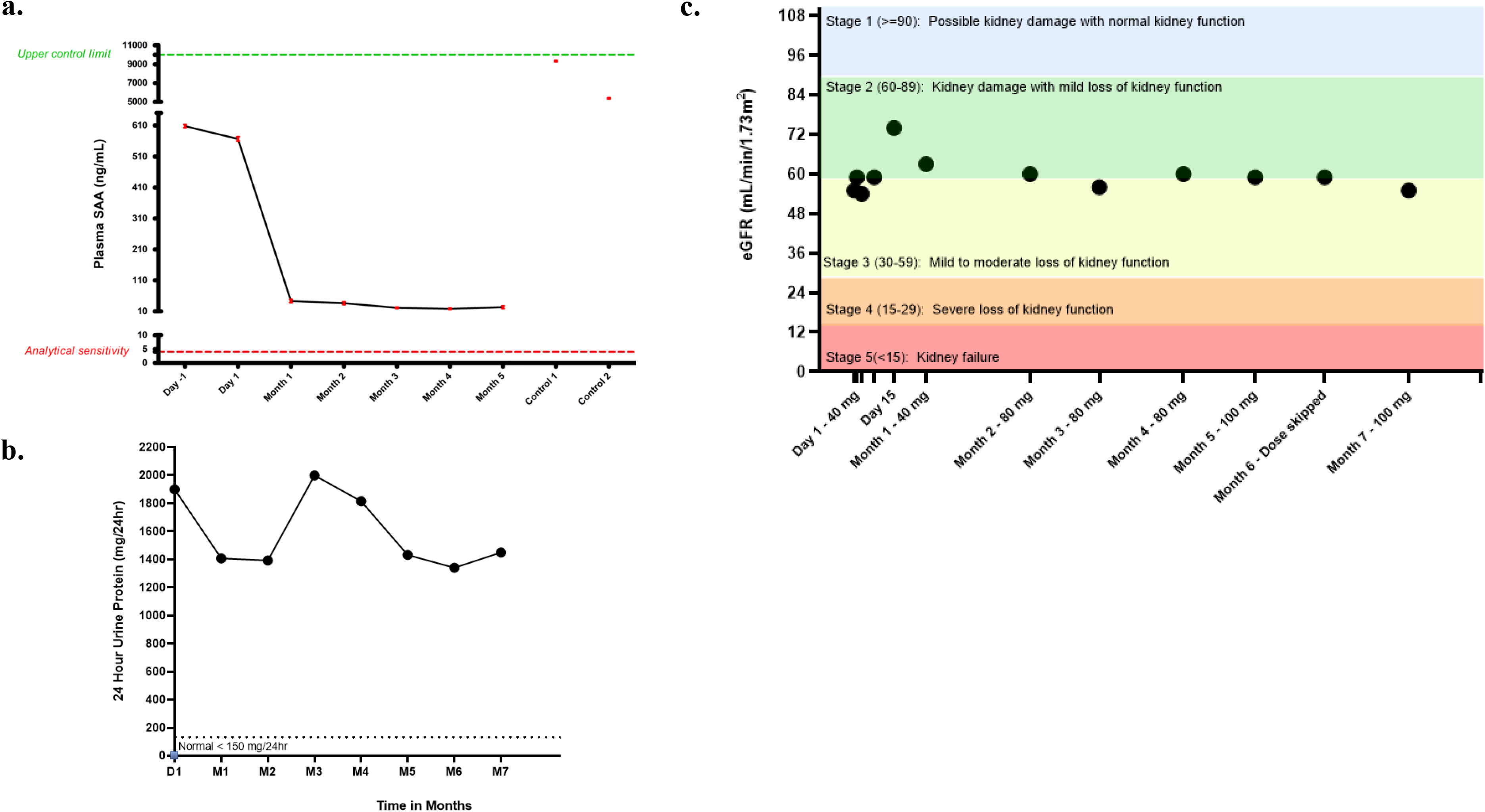
Renal performance parameters prior to and during treatment with experimental ASO treatment of in the SAA1 patient. a. Plasma SAA levels over time. The points represent means and standard errors of at least 3 replicate assays. b. The levels of protein in the patient’s urine over time. c. Glomerular filtration rate (GFR) over time. The urinary protein levels and GFR were determined using standard clinical tests. Plasma SAA measurements were as described in materials and methods.

### *SCN2A* Patient

A male child patient when treatment was initiated had a heterozygous c.5645G>A toxic gain of function mutation in the sodium voltage-gated channel alpha subunit 2 (*SCN2A*) gene. The *SCN2A* gene encodes the alpha subunit of Nav 1.2, a neuronal voltage-gated sodium channel [38–40][40–42]. Various types of pathological mutations have been reported throughout the *SCN2A* gene and the severity of disease appears to correlate with the type of mutation, with toxic gain of function mutations causing the most severe forms, namely infantile epileptic encephalopathy [40]. The patient presented with severe, intractable seizures at birth that have been quite consistent throughout life. Despite treatment with as many as 10 anti-epileptic medications, had consistently experienced 20-40 seizures per month, and had to receive rescue medication (nasal midazolam and/or rectal diazepam) weekly, to be admitted to the hospital to gain seizure control approximately twice yearly. At the initiation of treatment, in addition to severe, poorly controlled seizures, the patient was non-verbal, developmentally delayed and considered to suffer from autism spectrum disorder. The patient had an overall Adaptive Behavioral Composite score of 44 on the Vineland 3 Adaptive Behavior Scale (severe impairment) and age equivalent skills that ranged from 8 months to 5 years old with most in the 1-2 year age range. The patient struggled with irritability, behavioral problems, extreme sensory sensitivities and sensory seeking behaviors, as well as, intermittent pain and the manifestations of disease were progressing. Supplemental Table 6 shows the concomitant medications with which that patient was treated during ASO treatment. Importantly, the patient was treated with phenytoin and cenobamate, both sodium channel blocking agents and this substantially complicated ASO treatment and evaluation as the antisense medicine is designed to reduce the level of *SCN2A* encoded sodium channels. Since sodium channel activity must be tightly regulated and seizures can be caused by either excess or reduced sodium channel activity, as the ASO dose was increased, adjustment of the doses of the small molecule sodium channel blockers was required.

This patient requires an allele-selective ASO, as loss-of-function (LoF) mutations in *SCN2A* can lead to autism spectrum disorder and intellectual disability. Multiple non-pathogenic SNPs were identified in the patient’s *SCN2A* gene to which ASOs were designed, from which an ASO was selected as it was potent, highly allele-selective, and did not stimulate innate immunity (Table 6, Fig 3c). Nonclinical development study results are presented in Supplemental materials (Supplemental Material – Detailed methods and results for exemplary patients). The first IT dose was at 20 mg, followed by 40 mg one month later, and after a thorough assessment of safety and tolerability the dose was gradually escalated to 60 mg two months after the second dose, and lastly to 80 mg three month after the third dose. The patient has been treated for ∼8 months, with no ASO-related serious adverse events or adverse events, nor any emergent changes in the composition of the cerebrospinal fluid or other standard laboratory tests. The primary treatment goal is to reduce the frequency and severity of seizures, with a secondary goal to improve behavior and developmental delays. A significant, ASO-dose-dependent reduction in the frequency and severity of seizures (Fig. 4b) was observed. In fact, after the 40 mg dose, the patient was seizure free for a month for the first time in his life. However, as mentioned above, the concomitant administration of phenytoin and cenobamate complicate ASO treatment and assessment. After the 60 and 80 mg doses of ASO, the phenytoin was reduced as phenytoin is associated with significant adverse events. Even after the reduction of the phenytoin dose, significant improvement in seizure activity continued and the patient experienced short (up to 2 weeks) seizure free periods, with seizures appearing to increase toward the end of the dosing period. This is consistent with the known pharmakinetics/pharmacodynamics properties of IT administered ASOs and reflects a reduction in the concentration of the ASO in CNS tissues [15]. Dosing continues with the objective of identifying the optimal ASO dose and dose frequency and reducing or eliminating the administration of phenytoin.

Although formal assessment of cognitive and behavioral symptoms will be conducted after 1 year of ASO treatment, caregivers independently reported encouraging observations in sensory and repetitive behaviors. Specifically, one instance was reported where the patient was exhibiting less obsession about picking up a small object from the ground. Such behaviors prior to treatment were compulsive and repetitive.

### *SAA* Patient

A male patient who has biopsy-proven serum amyloid a (*SAA*) amyloidosis in the kidney. Genetic testing for familial periodic fever syndromes have been negative; however, a variant chr11: 18290751 A>T (hg19) encoding for a missense mutation in *SAA1* (NM_199161.5):c.101A>T(p.D34V) was identified in both siblings [47]. The mutation is thought to exacerbate the misfolding, precipitation, and aggregation of *SAA*. The patient has chronic kidney disease stage 3A with an estimated glomerular filtration rate (eGFR) of 52 ml/min/1.73 m^2^— losing 5-6 ml/min/1.73m^2^ per year for the last 2 years. The patients proteinuria was averaging 3.5 g/d but recently had decreased (Table 7). Renal biopsy demonstrated amyloid deposits. SAA levels were normal and no elevation of pro-inflammatory mediators was found.

**Table 7:**
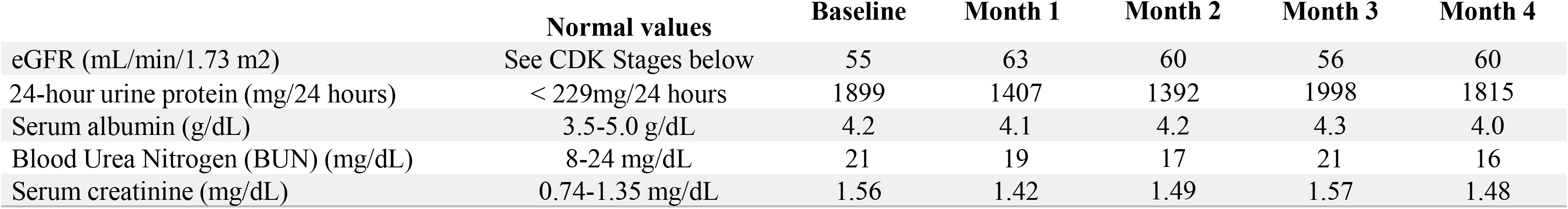
Laboratory analytes over time following treatment of SAA1 patient.

As the primary treatment goal is to reduce the rate of deterioration of renal function by reducing the plasma level of the non-essential protein SAA, non-allele-selective GalNAc PS 2′-MOE Gapmers were designed to hundreds of sites in *SAA* pre-mRNA and screened at 4000 nanomole in A431 cells. The most potent ASOs were evaluated in dose response experiments in the same cells (Fig. 3e), and the leads were then subjected to additional in silico and *in vitro* evaluations to identify potential off-targets and to exclude ASOs that are innate immune stimulatory and the optimal ASO selected (Table 6). Nonclinical development study results are presented in Supplemental materials (Supplemental Material – Detailed methods and results for exemplary patients) [38].

The primary analytes to be measured clinically are eGFR, serum creatinine, serum albumin, urinary protein levels and plasma SAA levels. Treatment was initiated SQ at 40 mg and has been dose-escalated to 80 mg to date. The patient has received the first four doses and values of key analytes are shown in Table 7. The first four doses have been well tolerated with no adverse events. Though there is insufficient data to definitively determine if the ASO is limiting further erosion of renal function, it is encouraging that after 4 months there is no decline in any of the relevant renal perimeters.

## DISCUSSION

Patients with extremely rare mutations constitute a tragedy of which we were largely unaware until the development of relatively inexpensive genomic sequencing. Though the scientific community is learning more about these individuals every day, our knowledge remains fragmented and primitive. For example, though the UDN has reported that average time from symptom onset to diagnosis in “undiagnosed” patients is perhaps as long as eight years [11], we now have sufficient data to report the median and mean times from symptom onset to diagnosis for patients expressing nano-rare mutations as 2 and 4.32 years, respectively. That the range was quite large, emphasizes how idiosyncratic the journey to diagnosis is for these patients. We suspect that numerous factors result in the enormous variation in time. Certainly, the age at which symptom onset occurs, the severity of the phenotype, the similarity of the phenotype to previously described syndromes and the organs affected are important considerations. However, circumstances such as proximity to a medical school, the level of research interest in the gene, the level of commitment of patients and parents to obtaining a diagnosis and the socio-economic level of the family affected play significant roles as well. Importantly, the patients who achieved a diagnosis and applied for treatment by n-Lorem are, we suspect, a tiny fraction of patients with ASO-targetable nano-rare mutations. Often, patients or parents persisted in pursuit of a diagnosis by contacting multiple physicians and scientists in various academic institutions and, in many cases, they had to pay for genomic sequencing and other basic research. Irrespective of the factors contributing to delays in diagnosis, the costs to society of this inefficiency are enormous, including those of care, the impact of incorrect diagnoses or inappropriate treatment and the loss in productivity of the affected families. Of similar concern is the inequity of the diagnostic process. Clearly, wealthier families have the means to pursue diagnosis and treatment that are unavailable to those less advantaged. Rather than discuss the various negative effects of the inequality of the delivery of health care, our position is simple: it is wrong.

We have also had several patients/parents who paid investigators to conduct the basic research necessary to make a genomic diagnosis and characterize the mutation who were unable to find a research physician at an academic center who was willing to treat the patient with an experimental ASO. This is despite the multiple personalized medicine centers being established. Tragically, this set of barriers to effective care could be eliminated by simply introducing genomic sequencing as a part of routine newborn evaluation protocols. Were this reform adopted, other tests currently performed to identify specific genetic diseases could likely be discontinued, reducing the net cost to the health care system.

At n-Lorem, though n-Lorem is fully independent of Ionis, because the lead author of this manuscript was the CEO and lead scientist at Ionis and several n-Lorem leaders were trained at Ionis, we have combined more than three decades of experience in antisense technology with decades of experience in drug development to establish systems that maximize quality at each step. The process begins with the complex and challenging risk/benefit evaluation necessary to assess which patients are appropriate candidates for personalized ASO treatment to the creation of novel approaches to assessing the performance of each experimental ASO and the aggregate benefit to patients achieved in an industrialized approach. Of particular importance is the challenge of identifying an optimal ASO for each patient. ASOs are complex bioactive molecules and the difference between an optimal vs sub-optimal ASO can be significant. We have published how we identify optimal ASOs and the procedures described are the product of having studied millions of ASOs and, in our view, represent the minimum acceptable process [9, 10]. We believe that if the processes we have published, including evaluating the recommended number of ASOs at each step, and the ASOs are dosed appropriately, severe adverse events (SAEs) should be rare, but should unexpected SAEs occur, it is vital that they must be managed professionally. This includes meeting regulatory requirements. For example, the FDA requires that reports be filed within 15 days of notification of the event. While notification of the FDA is mandated by regulations and vital, given the level of interest and the number of investigators and institutions focused on treating nano-rare patients with ASOs, it is also critically important to disseminate information about the SAE to all relevant audiences as rapidly as possible. Finally, particularly if the same ASO is to be provided to another patient, any new SAE should be included in an updated informed consent document.

To our knowledge, this is the first effort to correlate detailed genetic data with individual phenotypic observations in nano-rare patients. Though we report only a small number of these patients, the data are already sufficient to enable a variety of insights. We also believe that our effort is the first industrialized approach that couples detailed genotypic and phenotypic data to the immediate potential for treatment. We have demonstrated that the efficiency of antisense technology coupled to the special guidance for ASOs issued by the FDA makes it possible to mount an individualized ASO discovery program and move rapidly and inexpensively to treatment. This is an important step that should incentivize disease-focused patient advocacy groups to consider broadening their investment strategy to include grants to treat some patients today. This effort is enabled by decades of advances in basic research and in drug discovery technologies, demonstrating in a direct manner, the value of such investments. It is also important to note that only the combination of a novel, more efficient drug discovery platform, a novel non-profit model and the active commitment and cooperation of the FDA to help these desperate patients makes this possible. We believe there is potential value in considering what has been accomplished to date as a model that could help solve other challenging health delivery issues.

As mutations occur throughout the genome, it is as expected that pathogenic mutations were observed in diverse gene families and that genes that encode proteins with critical functions at key junctions in biological networks are over-represented and result in more diverse and severe phenotypes. Our experience is significantly biased by the preponderance of applications to treat patients with neurological diseases. Not surprisingly, in the CNS, mutations in ion channels, proteins involved in endosomal migration along axonal microtubules, and mutations in proteins involved in signaling, are well represented. Additionally, mutations in proteins involved in transcription, RNA processing and translation are associated with diverse serious illnesses. We also observe that mutations of all types are represented. We also extend the observation that, for more common rare diseases, the pattern of one or two common mutations accounting for most patients and a plethora of extremely rare mutations accounting for many other patients are common. That said, the frequency of the involvement of some gene families is notable. Mutations in genes that encode ion channels account for 26 patients in this cohort. This attests to the importance of ion channels in the CNS. One would expect mutations in ion channels to be significant contributors to diseases in any organ that must rhythmically depolarize and repolarize cell such as the heart. Though many of the ion channels in this cohort are primarily expressed in the CNS, it will be of interest to follow these patients to see if other organs are affected as the disease progresses. We also observe that many patients with mutations in ion channels experience gastrointestinal (GI) and genitourinary (GU) issues, posing the question whether some of these patients could have autonomic dysfunctions. If genomic sequencing were performed on all newborns, these patients would present remarkable opportunities to perform molecular epidemiological studies that could result in a step-by-step understanding of how a single mutation alters the molecular phenotype over time and answer why some patients experience earlier or later onset of symptoms, to say nothing of the opportunity to treat many of these patients early in their disease or pre-symptomatically.

While it is perhaps not too surprising that equivalent mutations in the same gene may result in phenotypes shared by patients with the same mutation and in manifestations that differ from the shared phenotype in some patients, and that some patients may display differences in phenotype. Clearly, our database supports a more thorough assessment of such differences and the opportunity to perform additional studies and follow up to better understand the molecular mechanisms that underly the clinical observations. Perhaps some differences in phenotypes may be explained by variations in the stage of the syndrome or the age or sex of the patients, but most cannot be explained in such a manner. Consequently, most of these unique patients present a remarkable opportunity to apply “modern omics” and other approaches to begin to understand the various homeostatic, compensatory and secondary effects of these mutations on the networks that result in expression of their unique phenotypes. Comparison of the “omics” of the unique patients to those who express only the shared phenotype could add substantial new insights. For those patients who are amenable to ASO treatment it will also be of interest to learn which elements of the phenotypes respond and in what order.

The 4 exemplary patients described above provide an opportunity to present (primarily in supplemental materials) the detailed data derived from ASO discovery and development activities performed by n-Lorem, collaborators and the network of clinical research organizations (CROs) we have established that we consider the minimum necessary to assure that each patient is treated with an optimal ASO. The specific patients selected epitomize the diversity of nano-rare patients and some of the challenges they present. Both the *KIF1A* and *SCN2A* patients required allele-selective RNase H1 ASOs. The identification of potent, safe allele-selective ASOs is more challenging than the process involved to provide non-allele selective ASOs. This requires the identification of non-pathogenic SNPs that can be used to design ASOs with potential allele selectivity. As the number of non-pathogenic SNPs varies and the more SNPs, the greater the likelihood of identifying an attractive ASO, while a limited number of SNPs may make the process less likely to succeed. To achieve allele selectivity, we screen multiple ASOs that “tile” the SNP and the primary reason allele selectivity can be achieved is the effect of the sequence of the ASO/RNA heteroduplex on the site specific cleavage pattern of RNase H1[49]. Narrowing the sequence space typically results in less potent ASOs and a greater propensity to identify innate immune active ASOs. Thus, the overall ASO discovery process takes more time and has a lower probability of success. The preliminary evidence of benefit with two allele-selective ASOs administered intrathecally is cautiously encouraging, but our experience is not extensive enough to begin to estimate an overall probability of success.

On the other hand, the *SAA* patient represented a straightforward opportunity from the antisense perspective, particularly as Ionis has two commercially approved ASOs that treat another form of renal amyloidosis, TTR amyloidosis. The family history puts in stark relief the damage done to families by many nano-rare mutations and the WGS results demonstrate a unique pathogenic variant that causes aggregation of SAA even in the absence of excess SAA production present in this family.

These patients also highlight the importance of the versatility of antisense technology, the complexity of nano-rare patients and the need to individuate therapeutic decisions based on the phenotype and needs of each patient. To be able to design ASOs to take advantage of different post-RNA binding mechanisms is vital to treating a broad range of mutations and molecular pathologic challenges. It is also important that PS ASOs are approximately equipotent in the cytosol and nucleus [50, 51]. To be able to administer PS ASOs by multiple routes of administration enhances the diversity of patients that can be treated and supports the option of local treatment such as in the eye or CNS which reduces the potential for systemic toxicities. Finally, to select different chemical classes of ASOs, such as GalNAc PS ASOs to selectively target RNAs in specific types of cells, such as hepatocytes, enhances both potency and safety. The therapeutic complexity of nano-rare patients also emphasizes the importance of senior judgement, the value of advisory groups such as the ATTC and the need to focus on the most significant needs of each patient. In traditional drug discovery and development, one focuses on populations of patient while at n-Lorem each decision must be made in the context of the individual patient.

This study has significant limitations, beginning with the relatively small number of patients represented, but we expect the number of patients to increase significantly in the coming years. An even greater limitation is the relatively short treatment periods in patients summarized in this manuscript. Obviously, this issue will be resolved in the next few years. Nonetheless, to have evidence of suggestive benefit in more than one patient is cautiously encouraging. There are also obvious selection biases. The relative preponderance of neurological disease does not, in our opinion, reflect the total population with nano-rare mutations. We ascribe this anomaly to the emotional impact of neurological diseases and consequent energetic patient advocacy groups, plus our inability to engage the inborn errors of metabolism, kidney and eye disease communities. We suspect that there also may be selection bias evident in the types of mutations submitted based on a broad understanding of the limitations of ASO technology. We hope to correct this imbalance in the near term by more effectively engaging these communities. Though we have confirmed many mutations and phenotypes, to a large extent we rely on the information provided by the research physicians who submitted the applications for treatment. Perhaps less apparent, but quite important is the challenge of acquiring consistent phenotypic data. Our data indicate that there is significant variability in the elements of phenotype that are reported depending on the particular interests of the treating physician. For example, in many of our patients with mutations in ion channels there are suggestions of autonomic dysfunction such as chronic GI distress, thermal regulation and nystagmus that are not identified in many applications. The wide variation in how terms used to describe phenotypes, severity, rates of progress and levels of disability make data analyses challenging. In the coming year, we plan to introduce standard terminology for phenotypes and other relevant parameters and questionnaires designed to prompt submitting physicians to consider element of phenotypes commonly overlooked.

This manuscript adds to the evidence that significant reforms in healthcare are needed if the needs of patients with extremely rare mutations are to be addressed in any meaningful way. Certainly, the most urgently needed reform is to introduce genomic sequencing into newborn evaluation protocols and to couple the genetic observations to long term epidemiological studies. However, if the maximum number of patients are to be treated today, insurers including Medicare and Medicaid must seize the opportunity to save lives and health care dollars by supporting diagnosis and treatment. The National Institutes of Health and other funding agencies need to work with patient advocacy groups to assure that the limited funds available for the care of nano-rare patients is used effectively to treat those who can be treated today, while simultaneously investing in research to identify more effective treatments in the future.

Importantly, this manuscript emphasizes that nano-rare patients are unique experiments of nature in which a single variable, a single mutation, causes a profound change in phenotype. The unique coupling of full genomic and phenotypic characterization with rapid intevervention with an ASO designed to affect the genetic cause of the syndromes presents an unparalleled opportunity to 1) better understand genotype-phenotype relationships, 2) how individual humans respond to mutations from the clinical to molecular level, and 3) how the effect of treatments that alter the expression of the cause of the syndrome affects the expression of various clinical manifestaions of the molecular pathological processes. Treatment of pediatric patients also provides the opportunity to assess the “plasticity” of the developing CNS, as developmental delays are arrested and, in some cases, reversed. Longer term, by combining analyses using various “omics” tools with genomic sequencing at birth, the opportunity to perform molecular epidemiological studies may provide novel insights into health and disease in human subjects.

## SUPPLEMENTAL FIGURES

**1. Optimal timeline for ASO drug discovery, development, and manufacturing.**

**2. Overview of Quality Processes**

## SUPPLEMENTAL TABLES

**1. Different mutations in the same gene.**

**2. Further detail of mutations from Table 2 relevant for phenotype assessment.**

**3. Mutations not amenable to ASO therapy.**

**4. Quality of life questionnaire for a KIF1A patient. Detailed assessment of the different domains overtime.**

**5. Clinical Severity and Improvement as assessed for a patient with KIF1A.**

**6. Concomitant medications for a SCN2A patient during experimental ASO treatment.**

## Investigator Network

All investigators who submitted the 173 applications that n-Lorem had received at the time of data cut off and evaluated are members of the ‘n-Lorem Investigator Network’ and are listed below.

**Abreu, Nicolas, M.D**., NYU Langone Medical Center

**Anderson, Eric, M.D.,** Rady Children’s Hospital, UC San Diego School of Medicine

**Anikster, Yair, M.D., Ph.D**., Sheba Medical Center

**Bain, Jennifer, M.D., Ph.D.,** Columbia University Irving Medical Center

**Baker, Joshua, D.O**., Ann & Robert H. Lurie Children’s Hospital of Chicago

**Bergin, Ann MarieMB, ScM, MRCP** Boston Children’s Hospital, Harvard Medical School

**Berry-Kravis, Liz, M.D., Ph.D.,** Rush University Medical Center

**Bhoj, Elizabeth, M.D., Ph.D.,** Children’s Hospital of Philadelphia (CHOP)

**Bock, Matthew, M.D.,** Rady Children’s Hospital-San Diego, Loma Linda University Pediatric Cardiology International Heart Institute

**Bonkowsky, Josh, M.D., Ph.D.,** University of Utah

**Borooah, Shyamanga, MBBS, MRCP (UK), MRCSEd, FRCOphth, Ph.D.,** Shiley Eye Institute, UCSD

**Butler, Ian, M.D.,** McGovern Medical School at The University of Texas Health Science Center, Houston (UTHealth)

**Calame, Daniel, M.D., Ph.D.,** Baylor College of Medicine

**Chandra, Bharatendu, MBBS**, University of Iowa Carver College of Medicine

**Chao, Hsiao-Tuan, M.D., Ph.D.,** Texas Children’s Hospital

**Chi, Ching-Shiang, M.D.,** Tung’s Taichung MetroHarbor Hospital

**Chung, Wendy, M.D., Ph.D.,** Boston Children’s Hospital, Harvard Medical School

**Costain, Gregory, M.D., Ph.D.,** Hospital for Sick Children, University of Toronto

**Cousin, Margot, Ph.D.,** Mayo Clinic Rochester

**Dagli, Aditi, M.D.,** Orlando Health Arnold Palmer Hospital for Children

**Dasouki, Majed, M.D.,** Advent Health Medical Group

**Day, John, M.D., Ph.D.,** Stanford School of Medicine

**Devinsky, Orrin, M.D.,** NYU Langone Medical Center

**Dickson, Patricia, M.D.,** Washington University School of Medicine

**Dowling, James, M.D., Ph.D.,** Hospital for Sick Children, University of Toronto

**Eichler, Florian, M.D.,** Mass General Research Institute, Harvard Medical School, Massachusetts General Hospital

**Evrony, Gilad, M.D., Ph.D.,** NYU Langone Medical Center

**Fink, John, M.D.,** University of Michigan Ann Arbor

**Friedman, Jennifer, M.D.,** Rady Children’s Hospital, UC San Diego

**Gertler, Tracy, M.D., Ph.D.,** Northwestern University Feinberg School of Medicine, Ann &

**Robert H. Lurie** Children’s Hospital of Chicago

**Gissen, Paul, Ph.D.,** UCL GOS Institute of Child Health

**Gleeson, Joseph, M.D.,** Rady Children’s Hospital, UC San Diego

**Gonzalez-Duarte, Alejandra, M.D.,** NYU Langone Medical Center

**Grunseich, Christopher, M.D.,** National Institute of Neurological Disorders and Stroke

**Helbig, Ingo, M.D.,** Children’s Hospital of Philadelphia (CHOP)

**Hirano, Michio, M.D**., Columbia University Irving Medical Center

**Hisama, Fuki, M.D.,** University of Washington

**Izumi, Kosuke, M.D., Ph.D.,** Children’s Hospital of Philadelphia (CHOP)

**Jen, Joanna, M.D., Ph.D.,** Integrative Headache Medicine of New York, Icahn School of Medicine at Mount Sinai

**Kaufmann, Horacio, M.D.,** NYU Langone Medical Center

**Kim-McManus, Olivia, M.D.,** Rady Children’s Hospital, UC San Diego

**Kimonis, Virginia, M.D.,** University of California-Irvine Medical Center

**Konersman, Chamindra, M.D.,** Rady Children’s Hospital

**Kuo, Sheng-Han, M.D.,** Columbia University Irving Medical Center

**Lanpher, Brendan, M.D.,** Mayo Clinic Rochester

**Leung, Nelson, M.D.,** Mayo Clinic Rochester

**McCourt, Emily, M.D.,** Children’s Hospital Colorado, University of Colorado

**McKeown, Sarah, M.S.,** Children’s Hospital of Philadelphia (CHOP)

**Mencacci, Niccolo, M.D., Ph.D.,** Northwestern University Feinberg School of Medicine

**Michelson, David, M.D.,** Loma Linda University Children’s Hospital

**Moufawad El Achkar, Christelle, M.D.,** Boston Children’s Hospital, Harvard Medical School

**Muriello, Michael, M.D.,** Medical College Wisconsin

**Ortiz-Gonzalez, Xilma, M.D., Ph.D.,** Children’s Hospital of Philadelphia (CHOP)

**Oskarsson, Bjorn, M.D.,** Mayo Clinic Jacksonville

**Poduri, Annapurna, M.D.,** Boston Children’s Hospital, Harvard Medical School

**Russell, Bianca, M.D.,** David Geffen School of Medicine at UCLA

**Saenz, Margarita, M.D.,** University of Colorado

**Shahani, Dave, M.D.,** Cook Children’s Health Care System

**Shneider, Neil, M.D.,** Ph.D., Columbia University Irving Medical Center

**Tekin, Mustafa, M.D.,** Miller School of Medicine, University of Miami

**Velinov, Milen, M.D., Ph.D.,** Rutgers Robert Wood Johnson School

**Yu, Timothy, M.D., Ph.D.,** Harvard Medical School

**Yukaitis, Christopher, M.D., Ph.D.,** Boston Children’s Hospital, Harvard Medical School

**Zaidman, Craig, M.D.,** Washington University School of Medicine

## Data Availability

All data produced in the present study are available upon reasonable request to the authors

## Acknowledgments

**Individuals:** The authors gratefully acknowledge Rosanne Crooke, Ph.D., Frank Bennett, Ph.D., for broad contributions to the advancement of n-Lorem and for the helpful editorial comments. Also, Kim Butler for outstanding administrative support and leadership in n-Lorem. We also acknowledge many others at n-Lorem who contribute specific analyses used in this manuscript.

## Sources of support for n-Lorem

n-Lorem is supported by contributions from many individuals and organizations, including Drs. Stanley and Rosanne Crooke. n-Lorem is a recipient of funding from Ultra-rare Gene-based Therapy Network (URGenT) grant 13799744 of the National Institute for Neurological Disorders and Stroke (NINDS) with the principal investigator being Dr. Neil Shneider of Columbia University.

## Institutions

The authors thank all of the physicians, patients, and families who have submitted applications to n-Lorem.

## Conflicts of Interest

All n-Lorem authors are compensated by n-Lorem. Dr. Crooke, Chairman and CEO of n-Lorem receives no compensation. As a non-profit n-Lorem does not provide stock options or RSUs.

## Ethics Committees / Institutional Review Boards

All patients treated in this manuscript were treated under FDA approved INDs and after approval from each Institutional Review Board(s) (IRB).

Ethics committee/IRB of Mayo Clinic Institutional Review Boards gave ethical approval in IRB Application # 22-005092 for this work

Ethics committee/IRB of Children’s Hospital Colorado gave ethical approval in IRB Application # 22-2313 for this work.

Ethics committee/IRB of Columbia University Medical Center gave ethical approval in IRB Application # AAAU1165 for this work.

Ethics committee/IRB of Rush University Medical Center gave ethical approval in IRB Application # irbm-006-002 for this work.

## Notes

### Competing Interest Statement

The authors have declared no competing interest.

### Clinical Trial

IND161670; IND162638; IND166588; IND159493

### Funding Statement

This study was funded by donations to n-Lorem.

### Author Declarations

Informed consent was obtained from all patients involved prior to study start. All patients treated in this manuscript were treated under FDA approved INDs and after approval from each Institutional Review Board(s) (IRB). Ethics committee/IRB of Mayo Clinic Institutional Review Boards gave ethical approval in IRB Application # 22-005092 for this work Ethics committee/IRB of Children's Hospital Colorado gave ethical approval in IRB Application # 22-2313 for this work. Ethics committee/IRB of Columbia University Medical Center gave ethical approval in IRB Application # AAAU1165 for this work. Ethics committee/IRB of Rush University Medical Center gave ethical approval in IRB Application # irbm-006-002 for this work.

### Summary of Updates

We have shorten the manuscript by deleting one of the exemplary patients, FLVCR1. Though this patient is of interest because the he/she has a retinal disease and is being dosed intravitreally, we have included the safety and tolerability data in our safety summary. As it is too early to determine if the patient has benefitted from treatment, we do not think this example merits inclusion. This shortens the manuscript quite a bit. We have also added a new Figure 1 that outlines the step-by-step process used to make these challenging risk/benefit decisions and this is discussed in detail in the revised manuscript and included a typical ATTC presentation (supplemental material).

