## Supplemental Materials for "Genotypic and phenotypic analysis of 173 patients with extremely rare pathogenic mutations who applied for experimental antisense oligonucleotide treatment"

**Supplemental Material – Detailed Methods and Results for Exemplary Patients**

### Patient nL00255-KIF1A

#### Nonclinical Methods

ASO design and *in vitro* screen

The ASO was designed to bind the reference sequence at positions hg38:Chr2: 240737833 to 240737853 which is in cis with the *KIF1A* pathogenic variant (Figure 3A and Table 6). This approach supports differential binding to the RNA by the ASO and reduction of the pre-mRNA and mRNA from the pathogenic allele, while sparing the pre-mRNA from the reference allele by RNase H1. The ASO is a PS/PO gapmer with the sequence and 2'substitutions noted and PS/PO content shown. The sequence and chemistry are as follows:

5′-AUoGoUoACATTTTCTTTGoUoUGC-3′ The underlined nucleotides contain 2' MOE modification, and 'o' indicates phosphodiester linkages. All other linkages are phosphorothioate. All cytosine bases are modified at the 5methyl C as are the uracil bases in the 2' MOE nucleotides. The ten nucleotides in the middle portion (Gap) that are not underlined all contain 2′-deoxyribose.

*In vitro* potency and allele-selectivity were determined following a dose response of nL-KIF1-001 in iPSC-derived neurons from the patient's fibroblasts using free uptake at concentrations of 0.6 µM, 2.5 µM, 10 µM, and 40 µM for 16 hours followed by 1-week maintenance in ASO-free media. Cells were harvested and RNA isolated using RNA easy 96 well kit at Qiagen (Valencia, CA) according to the manufacturer's instructions. The expression of each allele was then measured using tagged probes (wild type or reference transcript" tagged in green and "pathogenic transcript" tagged in red). *KIF1A* mRNA was quantified using quantitative RT-PCR (TaqMan) assay on an Applied Biosystems 7900H Fast Real-Time PCR system at ThermoFisher Scientific (Waltham, MA).

A bioinformatic analysis was performed to predict complementarity of the nL-KIF1-001 sequence to human putative primary transcripts. Human primary putative transcripts identified as partially complementary to nL-KIF1-001 with 1-, or 2-base mismatches, or 3-base mismatches with 17 matches in a row and expressed in the central nervous system were considered potential "off target" genes. These potential "off targets" effects were confirmed by PCR analysis in appropriate cell lines.

Finally, the potential for ASOs to activate innate immunity was tested in an *in vitro* assay using BJAB cells (EBV-negative, B lymphoma cell line, ACC 757) measuring expression levels of CCL22 as a surrogate measure of inflammatory response in BJAB cells and comparing with benchmark ASOs of known proinflammatory profile.

Toxicology studies

*8-week single ICV dose tolerability study with nL-KIF1-001 in mice*

Four C57/BL6 female mice/group were assigned to treatment groups receiving a single ICV injection of phosphate buffered saline or nL-KIF1-001 at 0.7 mg. Terminal sacrifice was performed on day 56. The assessment of tolerability was based on mortality, clinical observations, body weight, and acute and delayed functional observation evaluation. In addition, brain and thoracic spinal cord were evaluated by standard histopathology, along with microglial cell activation (Aif1 and Cd68 mRNA evaluation and IBA1 immunohistochemistry), astrocyte reactivity (GFAP mRNA evaluation), and neuronal degeneration (calbindin immunohistochemistry). This study was conducted at Ionis Pharmaceuticals Carlsbad, CA.

*8-week single IT dose tolerability study with nL-KIF1-001 in rats*

Four Sprague Dawley (Crl:CD(SD)) male rats/group were assigned to treatment groups receiving a single IT injection of PBS or nL-KIF1-001 at 3 mg. Terminal sacrifice was performed on day 56, and the assessment of tolerability was as described for mice. This study was conducted at Ionis Pharmaceuticals Carlsbad, CA.

*A 13-week repeat dose toxicology study of nL-KIF1-001 by IT injection in rats*

10 Sprague Dawley (Crl:CD(SD)) rats/sex/group were assigned to treatment groups receiving artificial CSF or nL-KIF1-001 at 0.3 and 1 mg/dose on days 1, 29, 57 and 85. Terminal sacrifice was performed on day 92, one week after the last dose on day 85.

The assessment of toxicity was based on mortality, clinical signs, body weights, food consumption, functional observation battery, ophthalmology, clinical pathology parameters (hematology, coagulation and clinical chemistry), gross necropsy findings, and histopathological examination of the brain, spinal cord, dorsal root ganglia, liver, kidney, heart and spleen. This study was conducted at Charles River Laboratories (Senneville, Canada).

Manufacturing and formulation of nL-KIF1-001

nL-KIF1-001 was manufactured by ChemGenes Corporation Wilmington, MA according to good manufacturing processes (GMP). As quality control, multiple release tests were performed, including tests for appearance (lyophilized white to off-white cake), identity (within 2 Da of expected mass of 7070.0 Da), identification (synthesis report matches expected sequence), purity (determined as 98.2% pure by high-performance liquid chromatography), moisture content (reported as 0.71%), sodium content (5.75%), bacterial endotoxin (0.032 EU/mg) pH (7.46), bioburden (total aerobic microbial count and total yeasts and mold counts less than 100 CFU/g each). nL-KIF1-001 was reconstituted in Elliott's B Solution and filtered through a 0.22 µm polyvinylidene fluoride membrane by the Columbia research pharmacy at concentrations required to meet dose level requirements. The dose was administered in a volume of 10 to 14 mL. Prior to dosing, the formulated nL-KIF1-001 was kept at room temperature for no longer than 4 hours.

### Clinical Methods

Study design

The ASO was administered intrathecally; the starting dose was 20 mg, and the dose was escalated by 20 mg at the discretion of the physician. To date, the actual dosing days and doses have been as follows: 20 mg on Days 1 and 31; 40 mg on Day 59; 60 mg on Days 115 and 206, then quarterly dosing at 60 mg-100 mg depending on the clinical effects observed.

Treatment goals and assessments

Prior to initiating treatment, treatment goals and the clinical tests to be evaluated were selected in consultation with the principal investigator, then baseline data were collected during the time the ASO was being developed. The primary treatment goal was to reduce the severity and frequency of the seizure disorder and was assessed by comparing the frequency and severity of seizures during the period immediately pretreatment to those parameters following treatment with the ASO. Seizure assessment included evaluation of the number of seizure spikes within overnight EEG and the number of average daily seizures recorded by the parent/caregiver via seizure diary. A minimum of 7 days of seizure count was collected after each dose.

The secondary treatment goal, to improve mobility by reducing spasticity, was assessed by comparing mobility and average number of falls recorded in the pre-treatment time period versus post-dose. Mobility was evaluated in the clinic via the 6-mintue walk test. The patient had to complete the test without ankle-foot orthosis (AFOs), i.e., barefoot, with the caregiver or physical therapist walking alongside the patient for safety. The number of falls was recorded each day by the parent/caregiver in a fall diary. Additionally, the parent/caregiver also noted if the patient used the wheelchair all day.

Effects on developmental delays and cognitive abilities were assessed throughout the study via the Differential Abilities Scales-II, a normed battery of cognitive and achievement tests that provides insight into how a child processes information.

A quality-of-life questionnaire was answered by the parent/caregiver; the questionnaire uses a 5-point Likert scale across 6 domains: Physical Health, Positive Emotions, Negative Emotions, Social Interaction, Leisure and Outdoors, and Independence. The answer choices on this scale ranged from "Never" to "Very often," with the responses being quantified on a linear scale from 0 to 100. The time period to consider for the assessment was the month prior. A comparison of these ratings of quality of life at two timepoints (Baseline, before dosing began (9/26/2022), and Clinic evaluation 6, after the 6th dose (8/14/2023)) indicate marked improvements across all 6 domains in less than a year of dosing.

Two additional assessments, the Gross Motor Function Measure and the 9-hole peg test, were added in the middle of the study, as the patient gained fine motor skills and was able to perform those tests. Those are to be assessed every 6 months moving forward.

Lastly, one year after treatment initiation, the parents and the physician were asked to rate the patient's disease severity at study start and after one year of treatment and to assess improvement after one year of treatment.

### Patient nL00333-SCN2A

### Nonclinical Methods

ASO design and *in vitro* screen

This patient requires an allele-selective PS ASO. Non-pathogenic single nucleotide variants throughout the gene were identified via long-read WGS sequencing. Phased VCF files from a whole-genome sequencing data provider are filtered to variants that are located within the canonical transcript of the pathogenic gene and in cis with the pathogenic variant. The filtered variants are used to reconstruct the pathogenic allele of the patient. ASOs are then designed to target each variant by tiling the surrounding sequence. n-Lorem's algorithm generates ASOs that are micro-walked 1 base pair (bp) over the SNP site and are excluded if they do not meet specific parameters for sequence motif, such as nucleotide content, mononucleotide repeats, GC dinucleotides, pro-inflammatory motifs, and others. Through this algorithm, each ASO is then evaluated against the reference genome (GrCh38), as well as its mRNA, to identify the potential to hybridize at sites other than the intended target (off-targets). Then 20-40 PS 2'MOE Gapmer ASOs were designed around each non-pathogenic SNV and these were evaluated for potency and allele-selectivity in patient derived cells (Figure 4a and Table 6). The sequence of this PS/PO MOE gapmer is: 5' - TGoCoCoAoACAATGTACAAoGoGGT – 3' The underlined nucleotides contain the MOE modification, and 'o' indicates phosphodiester linkages. All other linkages are phosphorothioate. All cytosine bases are modified at the 5-position with a methyl group, as are the uracil bases in the MOE nucleotides. The ten nucleotides in the middle portion that is not underlined all contain 2′-deoxyribose.

Manufacturing and formulation of nL-SCN2-001

nL-SCN2-001 was manufactured by Avecia Nitto Denko (Cincinnati, OH) as described above.

**Clinical Methods**

Treatment goals and assessments

The primary treatment goal, to reduce the frequency and/or severity of the seizures, is compared to a baseline as described above.

The secondary treatment goal, to improve behavior, cognitive ability and language are compared to baseline using a battery of different tests.

- Adaptive behavior: Vineland Adaptive Behavior Scale Version 3 (Vineland-3) change in Growth Scale Value (GSV) scores for all subdomains.
- Irritability and aberrant behavior: Aberrant Behavior Checklist – Community Edition, (ABC-C); Repetitive Behaviors Scale – Revised (RBS-R).
- Sensory dysfunction: Short Sensory Profile- 2 (SSP-2).
- Cognitive and functional abilities: Bayley Scales of Infant Development-4 (BSID-4).
- Language and communication: Observer-Reported Communication Ability (ORCA).

**Patient nL00068-SAA1**

**Nonclinical Methods**

ASO design and *in vitro* screen

More than 450 ASOs were initially designed as non-allele-selective, PS 2′-MOE GalNAc gapmers. The GalNAc moiety enhances delivery to hepatocytes via binding to ASPGR, as the main source of SAA1 protein is hepatocytes (Prakash et al., 2014). *In vitro* screening was performed as described above except that more than 20 lead unconjugated ASOs were then conjugated to GalNAc and evaluated in hepatocytes. The sequence of the ASO is:

5’ -ACCCATTGTGTACCCTCTCC-3' The underlined nucleotides contain the 2′MOE modification, and all linkages are phosphorothioate. All cytosine bases are modified at the 5-position with a methyl group, as are the uracil bases in the MOE nucleotides. The ten nucleotides in the middle portion that is not underlined all contain 2′-deoxyribose.

Toxicology studies

*4-week twice weekly SC administration tolerability study with unconjugated nL-SAA1-001*

Four mice/group were dosed with PBS or 50 mg/kg ASO twice weekly for 4 weeks. Terminal sacrifice was performed on day 39. The assessment of tolerability was based on mortality, clinical observations, body weight, serum chemistry parameters and liver, kidney and spleen histopathology evaluation. This study was conducted at Ionis Pharmaceuticals (Carlsbad, CA).

*5-week once weekly SC administration study with nL-SAA1-001 and mouse surrogate in CD-1 mice*

Six male animals/group were assigned to treatment groups receiving weekly SC doses of PBS (with a loading dose on Day 4), nL-SAA1-001 at 50 and 80 mg/kg/week. In addition, four male animals/group received ION-1095368, the mouse-specific *Saa1* ASO, at 1, 5, and 10 mg/kg/week. Terminal sacrifice was performed on Day 40, approximately 96 hours after final dosing on Day 36. The assessment of tolerability and toxicity was based on mortality, clinical observations, body weight, clinical pathology parameters, organ weight, and histopathology evaluation. This study was conducted at Ionis Pharmaceuticals (Carlsbad, CA).

*A 13-week repeat dose toxicology study of nL-SAA1-001 by SC injection in CD-1 mice*

Ten mice/sex/group were assigned to treatment groups receiving weekly SC doses of saline, nL-SAA1-001 at 10, 30 and 60 mg/kg/week, and ION-1095368, the mouse-specific *SAA1* ASO, at 10 mg/kg/week. Terminal sacrifice was performed on Day 94, approximately 48 hours after final dosing on Day 92.

The assessment of toxicity was based on mortality, clinical signs, body weights, food consumption, clinical pathology parameters (hematology, coagulation and clinical chemistry), gross necropsy findings, and histopathological examination. This study was conducted at the Korea Institute of Toxicology (Daejeon, Republic of Korea).

Manufacturing and formulation of nL-SAA1-001

nL-SAA1-001 was manufactured in a GMP facility by ChemGenes Corporation (Wilmington, MA) and was then formulated by Argonaut Manufacturing Services (Carlsbad, CA) as 1 mL 20 mg/mL ready to use sterile solution in PBS as described above).

### Clinical Methods

Treatment goals and assessments

The primary treatment goal to improve kidney function and delay the need for dialysis is assessed via laboratory tests of liver and kidney function, and via biomarker analysis of SAA protein and *SAA* mRNA comparing pre- to post-dose values. Performance status of the patients is assessed with the Eastern Cooperative Oncology Group Performance Status scale and quality of life is measured using the Linear Analog Scale Assessment (LASA). The patient was dosed monthly SQ with the disease shown in Table 7.

**Supplemental Materials - Results from Exemplary Patients**

**Patient nL00255-KIF1A**

### Nonclinical Results

*In vitro* results

The dose response analysis demonstrated an IC50 of 8 µM. There was no significant reduction of the mRNA level from the WT allele at any concentration tested, highlighting the high selectivity of nL-KIF1-001 for the pathogenic transcript over wildtype (Figure 3B). The lead ASO had no significant off target effects and was not innate immunostimulatory in the BJAB assay.

GLP Toxicology results

Observations in the GLP toxicology study were similar to those reported for other PS/PO 2′-MOE gapmers (Collotta D, et. al. Front Pharmacol. 2023; Henry, S.P. & Danis R.P., Expert Opinion on Pharmacotherapy 2001).

### Clinical Results

Safety

The safety profile of this ASO was unremarkable, with most adverse events being mild and not related to study drug.

Treatment goals

Treatment goals were achieved as discussed in the primary manuscript. These results were further supported by the parent narrative.

Parent narrative supporting benefits of the study drug:

- *Her tremor is almost gone, meaning she can now hold a baseball.*
- *She no longer falls because of her disease, now she may fall because she is so excited to do something.*
- *She sings.*
- *The pain in her hands and feet resolve with treatment but comes back close to the next dose; hopefully we can fine-tune the dose, so she does not have pain.*
- *She is able to make it through the day without a nap.*
- *In our KAND situation, 'moderately ill' in the scheme of average KAND patients is huge considering our daughter's prognosis and rate of regression at time at first dose. The elements of tremor, engagement, and the fact that she's out of her chair are epic, certainly not moderate, but other challenges of KAND (primarily worsening vision, reflux, etc.) that*

*weren't necessarily "targets" of the ASO, as we understood, are continuing to follow the course of disease. So those elements of disease offset the massive improvements in mobility, fine motor, speech, and independence that this ASO has so incredibly provided for her.* (This references their rating of "moderately ill" after one year on treatment on the Clinical Impression – Severity)

- *Every movement has more purpose.*

### Patient nL00333-SCN2A

#### Nonclinical Results

*In vitro* results

The dose response analysis showed that nL-SCN2-001 had an IC50 of 2.6 μM for the pathogenic transcript. Importantly, there was no significant effect noted on the wildtype mRNA levels and the allele-selectivity was estimated to be 42-fold for the pathogenic transcript over the wildtype (Figure 3C). The lead ASO was found to have no significant off targets and was negative for innate immune activation in the BJAB assay.

Toxicology studies

Results from the GLP toxicology study were consistent with previous reports for ASOs of this chemical class (Collatta et.al., Front Pharmacol. 2023; Henry, S.P., et.al., 2008).

#### Clinical Results

Safety

The safety profile of this ASO was unremarkable, with all adverse events being mild and not related to study drug.

Treatment goals

Treatment goals and results are discussed in the primary manuscript.

### Patient nL00068-SAA1

#### Nonclinical Results

*In vitro* results

The dose response analysis showed that the unconjugated version of nL-SAA1-001 had an IC50 of 0.15 µM in A431 cells (Figure 3E). Results of other *in vitro* tests were similar to the other exemplary ASOs.

Toxicology results

Results of toxicology studies were similar to those reported for other GalNAc conjugated PS ASOs (Collatta et.al., Front Pharmacol. 2023)

**Clinical Results**

Safety

The safety profile of this ASO has been unremarkable.

Treatment goals

The results of treatment goals are discussed in the primary manuscript.
