## Supplemental Tables for "Genotypic and phenotypic analysis of 173 patients with extremely rare pathogenic mutations who applied for experimental antisense oligonucleotide treatment"

### Supplemental Table 1 (1 of 8).

| Gene | Mutation | Functional Consequence | Sex | Presenting symptoms | Current phenotype | Shared Phenotype | Difference/unique phenotype |
|---|---|---|---|---|---|---|---|
| ***CACNA1A*** | c.4177 G>A, p.V1396M | gain-of-function | male | Status epilepticus, abnormal eyes movements, hypotonia | Intractable seizures with recurrent episodes of status epilepticus, movements disorders (action tremor, ataxia), recurrent episodes of paroxysmal tonic upgaze, hemiplegic migraine attacks, global developmental delay, hypotonia | Intractable seizures with recurrent episodes of status epilepticus, movements disorders (action tremor, ataxia), hemiplegic migraine attacks, global developmental delay, hypotonia - Disease onset with seizures | Episodes of paroxysmal tonic upgaze noticed at disease onset<br>More severe seizures |
| | | | female | Respiratory difficulties, seizures | Intractable seizures (focal onset with secondary generalization and apnea, frequent episodes of status epilepticus), episodes of hemiplegic migraine attacks, movement disorders (ataxia, action tremor), behavioral issues (autism spectrum disorder), severe global developmental delay, hypotonia | | More severe seizures |
| ***CACNA1A*** | c. 4034T>C, p.L1345P | gain-of-function/ loss-of-function | female | Hypotonia, macrocephaly, global developmental delay | Seizures, nystagmus, movement disorders (dystonia), hypotonia, global developmental delay, cortical visual impairment | Seizures, movement disorders, global developmental delay, hypotonia | Nystagmus, visual impairment, dystonia (no tremor or ataxia),<br>No seizures at onset, No hemiplegic migraine attacks |
| | c.5420T>C, p.V1807A | | female | hypotonia, global developmental delay | Hemiplegic migraine attacks, oculomotor abnormalities (nystagmus, divergent strabismus) and visual impairment, movement disorders (action tremor, ataxia), behavioral issues (ADHD), severe global developmental delay/intellectual disability , hypotonia, sleep disturbance/insomnia | Movements disorders (action tremor, ataxia), hemiplegic migraine attacks, global developmental delay, hypotonia - Disease onset with seizures. | Oculomotor abnormalities (nystagmus, divergent strabismus) with visual impairment, behavioral issues (ADHD), and sleep disturbance/insomnia,<br>No seizures |

### Supplemental Table 1 (2 of 8).

| Gene | Mutation | Functional Consequence | Sex | Time to diagnosis | Duration of disease/ from 1st symptom | Presenting symptoms | Current phenotype | Shared Phenotype | Difference/unique phenotype |
|---|---|---|---|---|---|---|---|---|---|
| ***GNAO1*** | c.607G>A, p.G203R | Dominant negative | male | / | 2.5 years | Seizures, hypotonia | Seizures, movement disorders (chorea, dystonia), global developmental delay, hypotonia | Seizures, movement disorders (chorea, dystonia), global developmental delay, hypotonia - Disease onset with seizures | nil |
| | | | female | 11 month | 1 year 9 months | Seizures | Intractable seizures, hypotonia, movement disorders (predominantly chorea, dystonia), global developmental delay, visual impairment, cortical atrophy on brain imaging | | visual impairment |
| ***GNAO1*** | c.626G>A, p.R209H | gain-of-function/ loss-of-function | male | / | ~ 6 years | movement disorder (dystonia) | Hyperkinetic movement disorder (primarily generalized dystonia and superimposed paroxysmal chorea), global developmental delay with relatively preserved receptive language function, Seizures. | Seizures, hyperkinetic movement disorders (dystonia, chorea), global developmental delay (predominantly motor delay) | onset with movement disorder (dystonia) |
| | | | female | 8 months | 9 months | Global developmental delay | Hypotonia, global developmental delay with delayed gross motor, fine motor and speech | Global developmental delay, hypotonia | No movement disorders, no seizures |

### Supplemental Table 1 (3 of 8).

| Gene | Mutation | Functional Consequence | Sex | Time to diagnosis | Duration of disease/ from 1st symptom | Presenting symptoms | Current phenotype | Shared Phenotype | Difference/unique phenotype |
|---|---|---|---|---|---|---|---|---|---|
| ***CACNA1A*** | c.4177 G>A, p.V1396M | gain-of-function | male | 4 months | 4 years | Status epilepticus, abnormal eyes movements, hypotonia | Intractable seizures with recurrent episodes of status epilepticus, movements disorders (action tremor, ataxia), recurrent episodes of paroxysmal tonic upgaze, hemiplegic migraine attacks, global developmental delay, hypotonia | Intractable seizures with recurrent episodes of status epilepticus, movements disorders (action tremor, ataxia), hemiplegic migraine attacks, global developmental delay, hypotonia - Disease onset with seizures | Episodes of paroxysmal tonic upgaze noticed at disease onset<br>More severe seizures |
| ***KCNQ2*** | c.908C>T, p.S303F | dominant negative/ gain-of-function | female | 2 month | 13 months | Seizures | Seizures (including infantile spasms, tonic, myoclonic, tonic-clonic and clonic seizures), global developmental delay, hypotonia and hypertonia, bilateral hip dysplasia, contracture of wrist with ulnar deviation and right arm dislocation, cortical visual impairment and optic nerve coloboma, GERD and laryngomalacia. Tongue and lip tie, food protein-induced enterocolitis syndrome. | Seizures, global developmental delay, abnormal tone (hypertonia and hypotonia), movement disorder (spasticity), cortical visual impairment. | Optic nerve coloboma, Musculoskeletal abnormalities (bilateral hip dysplasia, contracture of wrist with ulnar deviation and right arm dislocation), GERD and laryngomalacia, tongue and lip tie.<br>No movement disorder |
| | c.683A>G, p.H228R | dominant negative | female | < 1 month | 29 months | Seizures | Seizures, global developmental delay, hypertonia, movement disorder (spasticity), Autism spectrum disorder | | Behavioral issues (Autism spectrum disorder),<br>No ocular feature |
| | c.821C>T, p.T274M | | female | 7 weeks | 5 years | Seizures | Seizures, global developmental delay (non-ambulant, absent speech), hypertonia and hypotonia, movement disorder (spasticity) | | No ocular feature |
| | c.841G>A, p.G281R | gain-of-function/ loss-of-function | male | a few weeks | 2 years | Seizures and cyanosis | Intractable seizures (infantile spasms, tonic, and myoclonic seizures), movement disorders (dystonia, spasticity), severe global developmental delay, axial hypotonia and appendicular hypertonia, cortical visual impairment, gastrointestinal dysmotility with small bowel intussusceptions. MRI: significant supratentorial global white matter volume loss, including atrophy of the corpus callosum, optic nerves, hippocampi, and brainstem. | | Dystonia, gastrointestinal dysmotility with small bowel intussusception |

### Supplemental Table 1 (4 of 8).

| Gene | Mutation | Functional Consequence | Sex | Time to diagnosis | Duration of disease/ from 1st symptom | Presenting symptoms | Current phenotype | Shared Phenotype | Difference/unique phenotype |
|---|---|---|---|---|---|---|---|---|---|
| ***KIF1A*** | c.760C>T, p.R254W | dominant negative | female | / | ~ 9 months | / | Global developmental delay, movement disorders (spasticity and tremor), and hypotonia | Movement disorders, and global developmental delay/intellectual disability | Tremor, hypotonia |
| | c.914C>T, p.P305L | | female | ~ 1.5 years | ~ 5.5 years | Developmental delay | Seizures, developmental delay with regression/intellectual disability, visual impairment, movement disorder (spasticity), painful peripheral neuropathy.<br>Optic, cerebral, and cerebellar atrophies on imaging | | Seizures, visual impairment, peripheral neuropathy |
| ***MED13L*** | Exons 3-25 deletion | loss-of-function | female | ~ 2 years | ~12 years | Developmental delay | Global developmental delay (predominantly speech delay)/Intellectual disability, auditory processing disorder, visual impairment | Global developmental delay/intellectual disability, visual impairment, hearing issues | nil |
| | c.4609C>T, p.Q1537* | | female | 1 year | 5 years | Multiple congenital anomalies | Global developmental delay, visual impairment, congenital hearing loss, behavioral issues (ADHD), history of prematurity with neonatal hyperbilirubinemia, dysmorphic features (epicanthal fold, narrow palate), skeletal defects (metatarsus adductus, hand deformities, craniosynostosis) | | Behavioral issues (ADHD), dysmorphic features, skeletal defects, and history of prematurity |
| ***NALCN*** | c.1639A>G, p.M547V | dominant negative | male | 11 years | 14 years | Developmental delay | Intellectual disability, hypotonia, movement disorders (paroxysmal ataxia, mild dystonia), history of global developmental delay with regression.<br>Atrophy of the cerebellar vermis on imaging. | Developmental delay, hypotonia | movement disorders |
| | c.1798G>C, p.D600H | gain-of-function | female | 1 month | 7 months | multiple congenital anomalies | Hypotonia, developmental delay, skeletal defects (hip dysplasia, club foot, ulnar deviation), congenital heart disease (VSD, PFO), history of irregular breathing patterns | | Multiple congenital anomalies, irregular breathing patterns |

### Supplemental Table 1 (5 of 8).

| Gene | Mutation | Functional Consequence | Sex | Time to diagnosis | Duration of disease/ from 1st symptom | Presenting symptoms | Current phenotype | Shared Phenotype | Difference/unique phenotype |
|---|---|---|---|---|---|---|---|---|---|
| ***NPC1*** | c.1554-1009G>A and c.2702C>A, p.S901Y | loss-of-function | female | 5 years | 7 years | Neurological regression with deteriorating mobility and cognition | Infrequent seizures (~ 1 crisis per year), history of neurological regression with deteriorating mobility and cognition, movement disorder (progressive ataxia), muscle atrophy, hydrocephalus with persistent tental thickening on imaging, vertical supranuclear gaze palsy, history of brain hamartoma. No hepatosplenomegaly. | Neurodevelopmental regression, movement disorder (ataxia), history of severe global developmental delay, splenomegaly, and abnormal eye movements. | Hydrocephalus, muscle atrophy, history of brain hamartoma. No organomegaly |
| | c.3562G>T, p.E1188* and c.3754+1_3754+2dupGT | | female | ~ 3months | 7 years | Global developmental delay, splenomegaly, and eye movement disorders | Progressive motor dysfunction, dysarthria, abnormal eye movements, movement disorder (ataxia), history of severe global developmental delay, and splenomegaly | | dysarthria |
| ***PLA2G6*** | c.1613G>A, p.R538H and c.2370T>G, p.Y790* | unknown | female | ~2 years | 7 years | Developmental delay, hypotonia | Global developmental delay with regression, hypotonia, seizures, movement disorder (ataxia), nystagmus, dysphagia, and cerebellar atrophy. | Global developmental delay with regression, hypotonia, variable movement disorders (ataxia, dystonia, spasticity), dysphagia, nystagmus, and cerebellar atrophy. | Seizures |
| | c.386T>C, p.L389S and c.2035-926G>A | loss-of-function | male | < 1 year | 8 months | Developmental delay with delayed motor milestones noted | Global developmental delay with regression, generalized hypotonia, movement disorder (dystonia), esotropia, oral-pharyngeal dysphagia, and abnormal brain imaging | | Esotropia, no nystagmus |
| | c.208 C>T, p.R70* and c.208 C>T, p.Arg70* | | female | ~ 1 year | 4 years | Developmental delay | Global developmental delay with regression, cortical visual impairment, nystagmus, central hypotonia, and movement disorder (spasticity) - Cerebellar atrophy | | Cortical visual impairment |

### Supplemental Table 1 (6 of 8).

| Gene | Mutation | Functional Consequence | Sex | Time to diagnosis | Duration of disease/ from 1st symptom | Presenting symptoms | Current phenotype | Shared Phenotype | Difference/unique phenotype |
|---|---|---|---|---|---|---|---|---|---|
| *SCN2A* | c.2558G>A, p.R853Q | gain-of-function/ loss-of-function | male | 4 years | 10 years | Extreme irritability, inability to sleep, and feeding difficulties noted immediately after birth; failure to thrive at milestone | Intractable seizures (early onset evolving from infantile spasms to tonic-clonic, tonic and myoclonic crises), severe global developmental delay (non-ambulatory, absent speech)/intellectual disability, movement disorders (choreoathetosis predominantly, and dystonia), gastrointestinal motility disorders (intermittent abdominal pain and discomfort, fecal incontinence, diarrhea alternating with constipation, GERD), anorexia, and dysphagia, sensory processing disorder. History of abnormal sleep patterns, kidney stone, anxiety disorder. | Intractable seizures, global developmental delay (with absent speech), hypotonia, movement disorders \, sensory processing disorder, gastrointestinal motility disorders, and extreme irritability | choreoathetosis, dysphagia, anxiety, history of kidney stones |
| | c.5645G>A, p.R1882Q | gain-of-function | male | / | / | Neonatal seizures | Intractable Seizures, history of global developmental delay (able to walk, but absent speech) and hypotonia, movement disorder (dystonia), sensory processing disorder, extreme irritability, frequent pain, gastrointestinal distress | | nil |
| | c.3973-1G>A | loss-of-function | male | / | / | / | Global developmental delay (predominantly speech), behavioral issues (autism spectrum disorder) | | No irritability, no sensory processing disorder, no gastrointestinal features, no seizures, no movement disorders, no hypotonia |

### Supplemental Table 1 (7 of 8).

| Gene | Mutation | Functional Consequence | Sex | Time to diagnosis | Duration of disease/ from 1st symptom | Presenting symptoms | Current phenotype | Shared Phenotype | Difference/unique phenotype |
|---|---|---|---|---|---|---|---|---|---|
| *TARDBP* | c.881G>T, p.G294V | gain-of-function/ loss-of-function | male | 1 year | 1 year | arm weakness, cramping and fasciculations | Deceased | limbs weakness, limited mobility | nil |
| | c.1055A>C, p.N325S | | female | 0 year | 1 year | Upper limb weakness | Primary issue is with mobility and walking that is affected by bilateral foot drop. Notes some early changes but speech is relatively normal | | nil |
| | c.1169A>G, p.N390S | | female | 2 years | 2 year | Upper limb weakness | Primary issue is with mobility and walking - currently very limited using walker and aides | | nil |
| | c.1127G>A, p.G376D | | male | 4 years | 6 years | Weakness and atrophy in the right upper limb (proximal) with rapid diffusion to the contralateral side | Locked-in syndrome. Unable to move limbs. Eyes have very limited motility which does not allow communication | | Poor verbal communication |
| | c.1144G>A, p.A382T | | female | 5 years | 9 years | Upper limb weakness | No lower limb movement (wheelchair restricted). No longer able to verbally communicate. Continues with slow use of hands to eat and type | | Poor verbal communication |
| | c.943G>A, p.A315T | | female | 2 years | 2 years | Lower limb (foot and leg) weakness | Hips and lower limbs weakness, lower back stiffness | | nil |
| | c.1147A>G, p.I383V | | male | 6 years | 10 years | Lower limb weakness | Non-ambulatory, beginning to note hand weakness, tremor | | Tremor |
| | c.983C>T, p.A328V | | male | 0 year | 3 years | Slurred speech and right-hand weakness | Hands weakness, slurred speech, though still intelligible, slight atrophy of upper extremities - EMG: widespread motor neuron disease with active and chronic denervation in the cervical and lumbar regions. Chronic denervation in the tongue, | | Dysarthria/poor verbal communication |

### Supplemental Table 1 (8 of 8).

| Gene | Mutation | Functional Consequence | Sex | Time to diagnosis | Duration of disease/ from 1st symptom | Presenting symptoms | Current phenotype | Shared Phenotype | Difference/unique phenotype |
|---|---|---|---|---|---|---|---|---|---|
| ***TUBB4A*** | c.1163T>C, p.M388T | gain-of-function/dominant negative | male | / | / | Nystagmus, impaired mobility with poor head control | Hypotonia, movement disorders (ataxia, dystonia, spasticity), global developmental delay, oculomotor abnormalities (esotropia, nystagmus) with visual impairment - hypomyelinating leukodystrophy on brain imaging | Movement disorders, global developmental delay, leukodystrophy | Eye movement disorders (esotropia, nystagmus) with visual impairment, hypotonia |
| | c.774T>C, p.D249N | gain-of-function | female | / | / | Abnormal balance | Movement disorders (dystonia, spasticity), developmental delay (predominantly gross and fine motor delay - normal cognition), dysarthria, scoliosis, partial left hip subluxation - Leukodystrophy on imaging | | Dysarthria, musculoskeletal anomalies |

#### Supplemental Table 2 (1 of 7).

| Gene | Gene function | Mutation (nucleic acid) | Mutation (amino acid) | Functional type of mutation | # of cases | Proximal molecular cause |
|---|---|---|---|---|---|---|
| CACNA1A | Calcium voltage-gated channel subunit alpha1 A | c.4034T>C | p.L1345P | gain-of-function/loss-of-function | 1 | For other mutations in this gene, evidence links missense mutations to channel hyperfunction, but his mutation has not been studied, Recent evidence (e.g., Jiang et al., 2019) suggest both GoF and LoF mutations in CACNA1A cause developmental epileptic encephalopathies |
| | | c.5420T>C | p.V1807A | gain-of-function/loss-of-function | 1 | unknown, could be gain or loss of function |
| | | c.4177G>A | p.V1396M | gain-of-function | 2 | Alters a highly-conserved valine in the S3 Helix in Domain III, V1396M is a GoF mutation that increase calcium influx as monitored by whole-cell currents and facilitated current activation |
| CHCHD10 | mitochondrial inner membrane organizing system (MICOS) system | c.44_45delinsTT | p.R15L | gain-of-function/loss-of-function | 2 | toxic gain evidence is more recent-R15L CHCHD10 mutant patients exhibit CHCHD10+ aggregates in the spinal cord and R15L animal model has widespread, progressive axonal swellings, duplication in CHCHD10 was also found to result in a similar disease (Neil told us), but R15L human cells exhibit lower levels of CHCHD10 protein and 50% reduction in zebrafish causes motor neuron pathology, double KO mice exhibit similar phenotype as CHCHD10 mutant mice |
| | | c.44_45delinsTT | p.R15L | gain-of-function/loss-of-function | 1 | |
| CLCN7 | Primary chloride/proton antiporter on lysosomes | c.2144A>G | p.Y715C | gain-of-function | 1 | Increased outward currents through the channel, associated with enhanced lysosomal acidification, Accumulation of cytoplasmic vacuoles in diverse cell types |
| | | not provided | UNK | loss-of-function | 1 | The primary target cell is osteoclasts. Deep intronic mutation resulting in activation in activation of a cryptic splice site, The patient has an intronic mutation resulting in a novel splice site resulting in frameshifting. A splicing strategy may be a suitable strategy to address this patient's disease |
| CLN3 | lysosomal function | 1. partial gene deletion 2. c.569dupG | 1. UNK, 2. p.A191Sfs*45 | total loss-of-function | 1 | |
| | | 1. IVS6-IVS23del4tCTCA 2. IVS8-IVS10del (1.02kb) | 1. UNK, 2. UNK | total loss-of-function | 1 | |

#### Supplemental Table 2 (2 of 7).

| Gene | Gene function | Mutation (nucleic acid) | Mutation (amino acid) | Functional type of mutation | # of cases | Proximal molecular cause |
|---|---|---|---|---|---|---|
| COL6A2 | Anchor basement membrane to surrounding extracellular matrix. May also provide anti-apoptotic signals and anti-oxidant signals to myofibrils, Major connective tissue protein | c.2098G>A | p.G700S | dominant negative/loss-of-function | 1 | |
| | | c.1402C>T | p.R468X | dominant negative/loss-of-function | 1 | homozygous collagen 6A2 nonsense mutation (R468X) |
| GBE1 | glycogen branching enzyme | 1.c.986A>C; 2.Intronic c.2053-3358_2053-3350delInsTGTTTTTTACATGACAGGT | 1. p.Y329S, 2. UNK | loss-of-function | 3 | Both the point mutation (Y329S) and splice mutation carried by this patient cause strikingly reduced GBE1 levels/activity, Impaired branching of glycogen, leading to accumulation of polyglucosan bodies, which contain abnormally branched glycogen. |
| GNAO1 | G protein subunit alpha 01 | c.607G>A | p.G203R | dominant negative | 2 | lower concentration maximally lowers cAMP than WT as opposed to loss-of-function variants which have a much higher concentration needed to inhibit cAMP (LoF verified by lower expression of GNAO1) |
| | | c.625C>T | p.R209C | gain-of-function/loss-of-function | 1 | The Arg209Cys variant of the GαO subunit has been studied in cell lines, the G203R variant has been studied in transgenic mice as well. Gain-of-function mutations in GαO subunit are associated with movement disorders, while loss-of-function mutations are associated with epilepsy. It is thought that the Arg209Cys variant is a gain-of-function mutation. |
| | | c.626G>A, | p.R209H | gain-of-function/loss-of-function | 1 | |
| H3F3A | Histone variant H3.3 | c.137C>T | p.T46I | gain-of-function | 2 | The precise mechanism of action isn't clear, but the investigator doesn't believe LoF likely, Atomic modeling suggests disease-causing mutations in H3F3A interfere with intra-histone, or DNA-histone, interactions, Likely influences the formation, deposition or stability of H3.3-contiaining nucleosomes. Wide range of phenotypes and severity with H3F3A mutations, suggest each may have a unique impact on these processes |

### Supplemental Table 2 (3 of 7).

| Gene | Gene function | Mutation (nucleic acid) | Mutation (amino acid) | Functional type of mutation | # of cases | Proximal molecular cause |
|---|---|---|---|---|---|---|
| KCNQ2 | potassium voltage-gated channel subfamily Q member 2 | c.908C>T | p.S303F | dominant negative/gain-of function | 1 | The Ser303Phe mutation in KCNQ2 has not been reported before. It is noted as likely pathogenic, Likely based on presentation of patient and impact of other known KCNQ2 variants in this region of the protein, The mutation occurs in segment 6 of the protein required for channel activity |
| | | c.683A>G | p.H228R | dominant negative | 1 | Impact of genetic change on gene function: Mutation is found within the cytoplasmic loop region between the transmembrane 4 and 5 regions which form the pore. Thought to have dominant-negative activity for the multimeric complex, globally reducing current amplitudes (3 pore mutations) or by a depolarizing shift of the activation curve (2 voltage sensor mutations) decreasing potassium currents at the subthreshold level at which these channels are known to critically influence neuronal firing. One mutation significantly reduced surface expression |
| | | c.821C>T | p.T274M | dominant negative | 1 | Causes almost complete loss of function, but also has a dominant negative effect, AS the protein forms a heterotetramer, one mutated protein in the complex poisons the entire channel activity |
| | | c. 841G>A | p.G281R | gain-of-function/loss-of-function | 1 | electrophysiological evidence that the mutation poisons the M-current channel |
| KIF1A | Kinesin family member 1A | c.760C>T | p.R254W | dominant negative | 1 | The Arg254Trp mutation results in a decrease in binding affinity of KIF1A to microtubules. The mutations affect the velocity and force generation of the motor, but the primary defect seems to be a decrement in binding affinity (Lam et al. BioRxiv 09 19 20, Decreased axonal transport of synaptic vesicle precursors and other cargo resulting in mis-localization of cargo |
| | | c.914C>T | p.P305L | dominant negative | 1 | thought to act as a dominant negative resulting in mis-localization of kinesin cargo proteins. Mis-localization of cargo, in turn results in neuronal toxicity, decrease in binding affinity of KIF1A to microtubules. The mutations affect the velocity and force generation of the motor, but the primary defect seems to be a decrement in binding affinity |
| MED13L | Component of the mediator complex links gener al transcription factors at the TSS to more distal transcriptional regula tory machinery | Exons 3-25 deletion | UNK | loss-of-function | 1 | There are several theoretical antisense approaches which could be used to increase expression of the unaffected allele, such as uORF, and blocking a microRNA binding site. However, to the best of my knowledge there are no data supporting that either mechanism will modulate MED13L expression |
| | | c.4609C>T | p.Q1537X | loss-of-function | 1 | nonsense mutation, There are several theoretical antisense approaches which could be used to increase expression of the unaffected allele, such as uORF, and blocking a microRNA binding site. However, to the best of my knowledge there are no data supporting that either mechanism will modulate MED13L expression |

### Supplemental Table 2 (4 of 7).

| Gene | Gene function | Mutation (nucleic acid) | Mutation (amino acid) | Functional type of mutation | # of cases | Proximal molecular cause |
|---|---|---|---|---|---|---|
| NALCN | This gene encodes a voltage-independent, nonselective cation channel which belongs to a family of voltage-gated sodium and calcium channels | c.1639A>G | p.M547V | dominant negative | 1 | Establishment of the resting membrane potential, and therefore excitability, of electrically-excitable cells, confirmed gain of function NALCN variant Y578S conducts a higher Na+ background current than WT-expression was significantly lower in NALCN Y578S mutant than WT suggesting enhanced activity, not just expression change in cells, in c.elegans mutant was found to reduce locomotion |
| | | c.1798G>C | p.D600H | gain of function | 1 | Other mutations in NALCN that cause CLIFAHDD cluster in this same pore-lining transmembrane helices, In heterologous cell systems, CLIFAHDD mutations cause increased NALCN currents and impaired inactivation |
| NPC1 | NPC intracellular cholesterol transporter 1 | 1. c. 1554-1009G>A | p.S901Y + p.N222S | loss of function | 1 | c.1554-1009 G>A mutation inherited from her mother which creates a cryptic splice site and pseudoexon that puts the sequence out of frame |
| | | 1. c. 3562G>T 2. c. 3754+1_3754+2 dupGT | 1. p.E1188Ter, 2. UNK | loss of function | 1 | compound heterozygous c.3754+1_3754+2dupGT and E1188* pathogenic variant in NPC1. This previously reported pathogenic stop-gain mutations and the novel splice site variant, likely cause loss of NPC1 function. There is no way for ASO to correct a 'splice loss' mutation. No obvious ways to 'exon skip' with an ASO |
| PACS1 | PACS1-phosphofurin acidic cluster sorting protein 1 | c. 607C>T | p.R203W | dominant negative/gain of function | 2 | The mutation causes aggregation of GFP-tagged PACS1 in human cells, enhances protein stability and decreases interaction with TRPV4 |
| PACS2 | Multifunctional sorting protein that controls the endoplasmic reticulum (ER)-mitochondria communication | c.625G>A | p.E209K | gain of function/dominant negative | 2 | Mutation in the unstructured middle region (MR), in a proposed autoregulatory domain, Altered interactions with client proteins leading to abnormal intracellular trafficking of, e.g., SIRT1, HDAC1, TRPV1 |
| PLA2G6 | Calcium-independent phospholipase A2, which preferentially cleaves the sn-2 acyl bond of phospholipids | creation of 147bp pseudo-exon | UNK | UNK | 1 | Deep intronic mutation creates a novel splice acceptor site, which reportedly results in creation of 147bp pseudo-exon with a PTC, driving nonsense-mediated decay, Loss of Phospholipase A2 mediated phospholipid remodeling, with consequences for membrane homeostasis, mitochondrial integrity and signal transduction |
| | | 1. c. 386 T>C 2. c. 2035-926 G>A | 1. p.L389S, 2. UNK | loss of function | 1 | |
| | | Homozygous c.208 C>T | p.R70Ter | loss of function | 1 | homozygous mutation is a truncation mutation in the second exon in the gene. The specific nucleotide change is adjacent to the 3′ splice donor site for exon 2 which may also impact splicing of exon 2, Loss of arachidonic acid signaling and accumulation of phosphatidylcholine in membranes. |

#### Supplemental Table 2 (5 of 7).

| Gene | Gene function | Mutation (nucleic acid) | Mutation (amino acid) | Functional type of mutation | # of cases | Proximal molecular cause |
|---|---|---|---|---|---|---|
| PLP1 | transmembrane proteolipid protein that is the predominant component of myelin. The encoded protein may play a role in the compaction, stabilization, and maintenance of myelin sheaths, as well as in oligodendrocyte development and axonal survival | | | gain of function | 1 | |
| | | Not provided | UNK | gain of function | 1 | |
| PPP2R5D | One of many B regulatory subunit of protein phosphatase 2A (PP2A) subunits | c.598G>A | p.E200K | dominant negative | 1 | Most disease-causing mutations in PPP2R5D, including p.E200K, occur in a conserved acidic loop which bridges the catalytic and scaffolding subunits of PP2A. (Houge et. al. 2015) propose a dominant-negative mode of action , Reduces interactions between the regulatory (B) subunit and the catalytic (C) and scaffolding (A) subunits of PP2A. |
| | | c.1258G>A | p.E420K | dominant negative | 1 | Strong - impact of E420K mutation in PPP2R5D has been mapped near the active site and is predicted to act in a dominant fashion (Shang et al., 2016), heterozygous E420K PPP2R5D in HEK cells show dominant effect |
| SAA1 | Serum amyloid A (SAA) proteins are apolipoproteins associate-ed with high-density lipoprotein (HDL) in plasma | none identified | UNK | gain of function | 1 | increase in SAA1 leads to amyloidosis due to production in the liver |
| | | none identified | UNK | gain of function | 1 | |

### Supplemental Table 2 (6 of 7).

| Gene | Gene function | Mutation (nucleic acid) | Mutation (amino acid) | Functional type of mutation | # of cases | Proximal molecular cause |
|---|---|---|---|---|---|---|
| SCN2A | voltage-gated sodium channel Nav 1.2 | c.2558G>A | p.R853Q | gain-of-function/loss-of-function | 1 | loss-of-function attributed to reduced electrical firing, gain of function with unregulated proton conductance |
| | | c.5645G>A | p.R1882Q | gain-of- function | 1 | Hyperexcitability of neurons thanks to increased sodium currents |
| | | c.3973-1G>A | UNK | loss-of- function | 1 | The patient has G to A mutation in the -1 position of the splice acceptor site for exon 22, resulting in skipping of exons 22-25; e.g., splicing of exon 21 to exon 26. The G residue at the -1-splice site is a critical residue, It is unlikely that an ASO will force proper splicing of exon 22. It may be possible to design an ASO that skip additional exons to make a truncated protein missing several exons. |
| TARDBP | TDP-43 (TARDBP) is an RNA and DNA binding protein involved in modulating transcription, RNA splicing, microRNA processing and DNA repair | c.881G>T | p.G294V | gain-of-function/loss-of-function | 1 | A hyperphosphorylated and cleaved form of TDP-43 is the major protein observed in ubiquitin positive, tau and alpha-synuclein negative protein aggregates observed in FTLD and ALS. Normally, TDP-43 is observed in the nucleus, however in ALS and FTLD tissue, TDP-43 is depleted from the nucleus, localized in cytoplasmic protein aggregates, This is a previously documented pathogenic mutation in the low complexity C-termini of TDP 43 causing loss of TDP-43 from the nucleus and toxic cytoplasmic aggregates |
| | | c.1055A>C | p.N325S | gain-of-function/loss-of-function | 1 | |
| | | c.1169A>G | p.N390S | gain-of-function/loss-of-function | 1 | |
| | | c.1135T>C | p.S379P | gain-of-function/loss-of-function | 1 | |
| | | c.1127G>A | p.G376D | gain-of-function/loss-of-function | 1 | |
| | | c.1144G>A | p.A382T | gain-of-function/loss-of-function | 1 | |
| | | c.943G>A | p.A315T | gain-of-function/loss-of-function | 1 | |
| | | c.1147A>G | p.I383V | gain-of-function/loss-of-function | 1 | |
| | | c.983C>T | p.A328V | gain-of-function/loss-of-function | 1 | |

#### Supplemental Table 2 (7 of 7).

| Gene | Gene function | Mutation (nucleic acid) | Mutation (amino acid) | Functional type of mutation | # of cases | Proximal molecular cause |
|---|---|---|---|---|---|---|
| TUBB4A | tubulin Beta 4A Class IVa | c.1163T>C | p.M388T | gain of function/dominant negative | 1 | OE of mutant TUBB4A in SHSY5Y cells suggests gain-of-function in which it caused hypomyelination phenotypes and altered ability of WT tubulin to incorporate into microtubules and decreased polymerization, which is further supported by KO mice and loss-of-function mutations, which increase polymerization. |
| | | c.774T>C | p.D249N | gain of function | 1 | Altered microtubule dynamics, which have been observed in both human cell lines with this mutation (Curiel et al., 2017) and knock in mouse models (Sase et al., 2020), Altered microtubule dynamics, and neurite outgrowth (Watanabe et al., 2017; Curiel et al., 2017; Vulinovic et al., 2018; Sase et al., 2020), KI mouse available (Tubb4aD249N/D249N),and have progressive neurodegenerative phenotype (Sase et al., 2020) |
| UBTF | A member of the HMG-box DNA-binding protein family | c.628G>A | p.E210K | gain of function | 3 | The mutation appears to be a gain-of-function mutation which markedly increases ribosomal RNA biosynthesis, Clear evidence that E210K mutation in UBTF creates a hyper-active UBF complex, leading to excess transcription of rDNA, Increased size and number of nucleoli and concomitant sensitivity to DNA damage, and cell cycle alterations |

**Supplemental Table 3.**

| Gene | Consequence | Acceptance Status | Explanation for Decline |
|---|---|---|---|
| CACNA1A | Gain of function/loss of function | Declined | Haploinsufficiency for this same gene leads to Episodic ataxia type 2 (EA 2), Phenotype of variant not adequately defined to be sure of strategy |
| CLCN7 | Loss of function | Declined | Target tissue not sensitive to ASOs, osteoclasts |
| CLN3 | Total loss of function | Declined | Too-large prevalence, Ionis drug discovery program |
| COL6A2 | Dominant negative/loss of function | Declined | Target tissue not sensitive to ASOs and unlikely correct molecular defect |
| MED13L | Loss of function | Declined | No antisense strategy |
| NPC1 | Loss of function | Declined | This previously reported pathogenic stop-gain mutations and the novel splice site variant, likely cause loss of NPC1 function. There is no way for ASO to correct a 'splice loss' mutation. No obvious ways to 'exon skip' with an ASO. |
| PLA2G6 | Loss of function | Declined | Technically infeasible |
| PLP1 | Gain of function | Declined | Prevalence >30 worldwide |
| SCN2A | Loss of function | Declined | Prevalence >30 worldwide |

### Supplemental Table 4.

| Quality of Life Questionnaire for KIF1A Patient | Baseline | Dose 1 | Dose 2 | Dose 3 | Dose 4 | Dose 5 | Dose 6 | Dose 7 |
|---|---|---|---|---|---|---|---|---|
| Date | 9/26/2022 | 10/26/2022 | 11/21/2022 | 1/18/2023 | 3/20/2023 | 5/20/2023 | 8/14/2023 | 11/8/2023 |
| Health and Well-being | | | | | | | | |
| Had enough energy to participate in daily routines and activities | Sometimes | Sometimes | Sometimes | Sometimes | Sometimes | Often | Often | Often |
| Kept in good general health (e.g., avoided coughs, colds, fever) | Sometimes | Sometimes | Sometimes | Sometimes | Often | Sometimes | Often | Often |
| Slept well during the night | Often | Often | Often | Rarely | Sometimes | Sometimes | Often | - |
| Been alert and aware during the day | Sometimes | Sometimes | Sometimes | Sometimes | Often | Sometimes | Often | Very Often |
| Feelings and Emotions | | | | | | | | |
| Been in a good mood | Sometimes | Sometimes | Sometimes | Often | Often | Often | Very often | Very often |
| Smiled or brightened their facial expression | Sometimes | Often | Often | Very often | Often | Often | Very often | Very often |
| Showed happiness through body language (e.g., making eye contact, body facing others) | Often | Often | Often | Very often | Often | Often | Very often | Very often |
| Showed cheeky or comical mannerisms (e.g., laughed, giggled) | Very often | Very often | Very often | Very often | Very often | Very often | Very often | Very often |
| Been unsettled without an apparent reason | Very often | Very often | Often | Very often | Very often | Very often | Often | Sometimes |
| Showed aggression (e.g., hitting, kicking, using offensive language, being destructive) | Rarely | Rarely | Rarely | Sometimes | Sometimes | Sometimes | Rarely | Never |
| Appeared upset or angry (e.g., crying, screaming, moving or stiffening the body) | Often | Often | Often | Sometimes | Very often | Very often | Rarely | Sometimes |
| Become withdrawn with a low mood | Often | Often | Sometimes | Sometimes | Rarely | Rarely | Rarely | Sometimes |
| Deliberately hurt themselves | Never | Never | Never | Never | Never | Never | Never | Never |
| Expressed discomfort with changes in routine (e.g., carers, school, out-of-home care) | Sometimes | Sometimes | Sometimes | Rarely | Never | Never | Rarely | Rarely |
| Showed signs of being anxious or agitated (e.g., teeth grinding, fast breathing, avoidance) | Often | Often | Sometimes | Sometimes | Sometimes | Sometimes | Rarely | Rarely |
| Caregivers | | | | | | | | |
| Expressed happiness when they were understood | Often | Often | Often | Very often | Often | Often | Very often | Very often |
| Appeared relaxed when making eye contact | Very often | Very often | Very often | Very often | Very often | Very often | Very often | Very often |
| Initiated greetings with people verbally or nonverbally (e.g., eye contact) | Often | Often | Often | Very often | Very often | Very often | Very often | Very often |
| Enjoyed being included | Often | Often | Often | Very often | Often | Often | Very often | Often |
| Enjoyed the social experiences of mealtimes | Rarely | Rarely | Sometimes | Often | Very often | Very often | Very often | Often |
| Responded positively when others paid attention to them (e.g., your child smiled, showed interest) | Often | Often | Often | Often | Very often | Very often | Very often | Often |
| Showed pleasure or excitement when looking forward to activities (e.g., going to school, outings, events) | Sometimes | Sometimes | Sometimes | Often | Very often | Very often | Very often | Often |
| Activities and the Outdoors | | | | | | | | |
| Enjoyed moving their body (e.g., walking, swinging, swimming) | Rarely | Rarely | Sometimes | Sometimes | Sometimes | Often | Very often | Often |
| Enjoyed feeling steady or stable during physical activities (e.g., sitting, standing, bike riding) | Rarely | Rarely | Rarely | Sometimes | Sometimes | Sometimes | Often | Sometimes |
| Enjoyed physical activities (e.g., going out for a walk, swimming, swinging, dancing) | Rarely | Rarely | Sometimes | Sometimes | Sometimes | Sometimes | Often | Sometimes |
| Enjoyed going on outings in the community (e.g., shopping, party, sports, theatre) | Sometimes | Sometimes | Sometimes | Sometimes | Often | Often | Often | Often |
| Enjoyed spending time outdoors (e.g., contact with water, grass, wind, sunshine) | Rarely | Rarely | Rarely | Sometimes | Sometimes | Sometimes | Very often | Sometimes |
| Daily Life | | | | | | | | |
| Expressed their needs (e.g., hunger, thirst, toileting) | Sometimes | Sometimes | Often | Often | Sometimes | Often | Very often | Very often |
| Made their own choices for activities or things they enjoy (e.g., DVDs, toys) | Often | Often | Often | Often | Often | Often | Often | Very often |
| Helped to complete routine activities (e.g., dressing, feeding, chores around the house) | Never | Never | Rarely | Rarely | Sometimes | Sometimes | Sometimes | Sometimes |
| Enjoyed making things with their hands - can be with help (e.g., building blocks, painting, cooking) | Often | Often | Often | Often | Often | Often | Often | Often |
| Enjoyed using technology (e.g., computer, tablet, applications on phones) | Often | Often | Often | Very often | Very often | Very often | Often | Often |

### Supplemental Table 5.

**Clinical Severity and Improvement as assessed by parents and physician for a KIF1A patient**

| Global assessment | Parent score | Physician score |
|---|---|---|
| **CGI-Severity at study start (scale of 0 to 7)** | 6 – Severely ill | 5 – Markedly ill |
| **CGI-Severity after 1 year of treatment** | 4 – Moderately ill | 4 – Moderately ill |
| **CGI- Improvement after 1 year of treatment compared to baseline (scale of 0 to 7)** | 2- Much improved | 2 – Much improved |

Question posed for the Clinical Global Impressions – Severity: Considering your total clinical experience with this patient population, how ill is the patient at this time?
Question posed for the Clinical Global Impressions – Improvement: Rate total improvement whether or not, in your judgment, it is due entirely to drug treatment. Compared to his/her condition at admission to the project (screening) how much has he /she changed?

**Supplemental Table 6. Concomitant medications for a SCN2A patient while on ASO treatment.**

| Medication Name (Generic name) | Indication | Dose | Frequency | Route |
|---|---|---|---|---|
| Phenytoin | Epileptic seizure | 5-200 mg | QD | PO |
| Midazolam | Epilepsy | 5 mg | seizure rescue medication (PRN) | Intranasal |
| Diazepam | Epilepsy | 10 mg | seizure rescue medication (PRN) | PO |
| Clonazepam | Epilepsy | 0.25 mg | seizure rescue medication (PRN) | PO |
| Cenobamate | Epilepsy | 150 mg | QD | PO |
| Melatonin | Insomnia | 0.25 mg | PRN | PO |
| Acetaminophen- and ibuprofen-containing product | Pain management | 100 mg | PRN | PO |
| Cephalexin | Bacterial infection | 250 mg | QID | PO |
