## Supplementary figures and images for "Genotypic and phenotypic analysis of 173 patients with extremely rare pathogenic mutations who applied for experimental antisense oligonucleotide treatment"

### Supplemental Figures

#### Supplemental Figure 1.

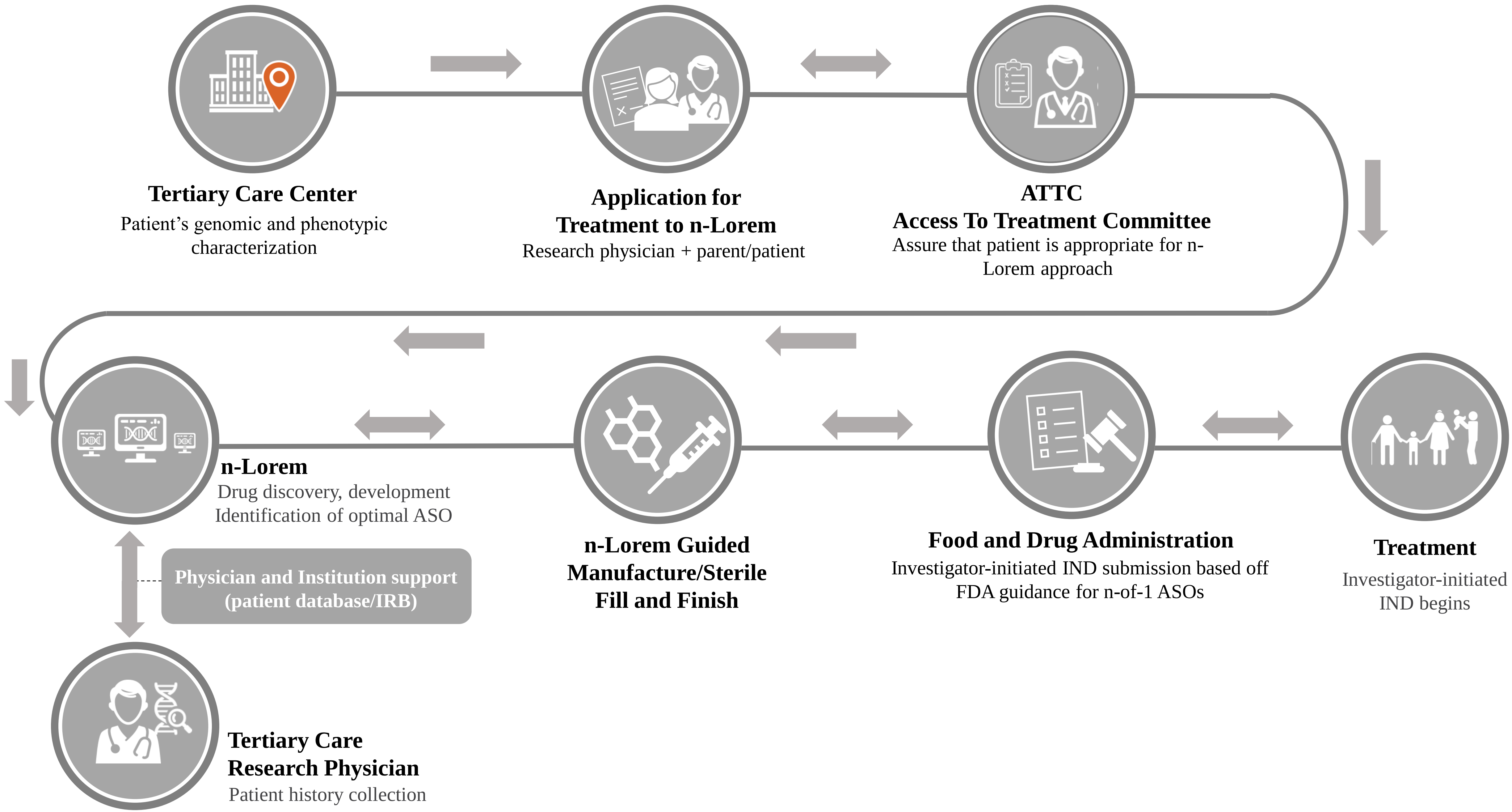

##### Supplemental Figure 2.

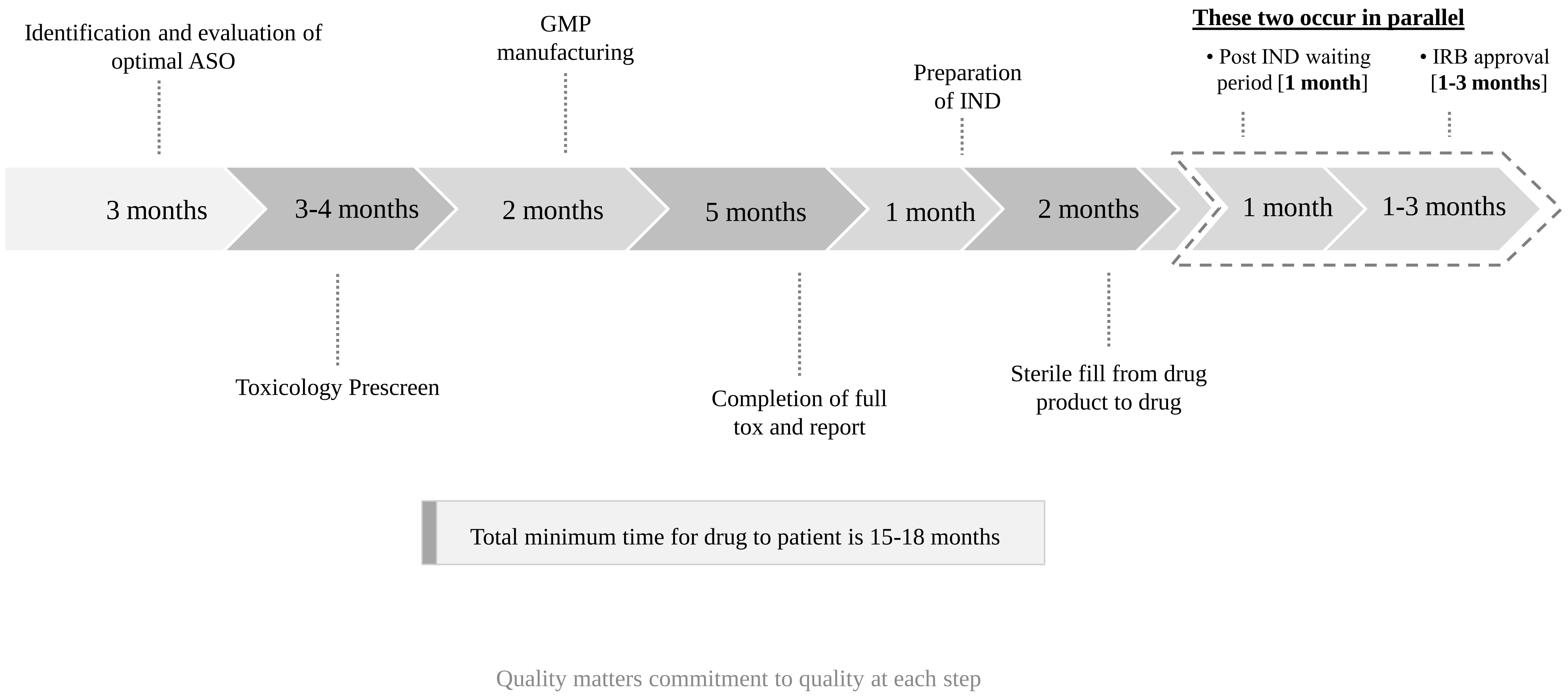
